# Environmental Exposure to Volatile Organic Compounds is Associated with Endothelial Injury

**DOI:** 10.1101/2021.08.25.21262556

**Authors:** Daniel W. Riggs, Marina V. Malovichko, Hong Gao, Katlyn E. McGraw, Breandon S. Taylor, Tatiana Krivokhizhina, Shesh N. Rai, Rachel J. Keith, Aruni Bhatnagar, Sanjay Srivastava

## Abstract

**Objective:** Volatile organic compounds (VOCs) are airborne toxicants abundant in outdoor and indoor air. High levels of VOCs are also present at various Superfund and other hazardous waste sites; however, little is known about the cardiovascular effects of VOCs. We hypothesized that ambient exposure to VOCs exacerbate cardiovascular disease (CVD) risk by depleting circulating angiogenic cells (CACs).

**Approach and Results:** In this cross-sectional study, we recruited 603 participants with low-to-high CVD risk and measured 15 subpopulations of CACs by flow cytometry and 16 urinary metabolites of 12 VOCs by LC/MS/MS. Associations between CAC and VOC metabolite levels were examined using generalized linear models in the total sample, and separately in non-smokers. In single pollutant models, metabolites of ethylbenzene/styrene and xylene, were negatively associated with CAC levels in both the total sample, and in non-smokers. The metabolite of acrylonitrile was negatively associated with CD45^dim^/CD146^+^/CD34^+^/AC133^+^ cells and CD45^+^/CD146^+^/AC133^+^, and the toluene metabolite with AC133^+^ cells. In analysis of non-smokers (n=375), multipollutant models showed a negative association with metabolites of ethylbenzene/styrene, benzene, and xylene with CD45^dim^/CD146^+^/CD34^+^ cells, independent of other VOC metabolite levels. Cumulative VOC risk score showed a strong negative association with CD45^dim^/CD146^+^/CD34^+^ cells, suggesting that total VOC exposure has a cumulative effect on pro-angiogenic cells. We found a non-linear relationship for benzene, which showed an increase in CAC levels at low, but depletion at higher levels of exposure. Sex and race, hypertension, and diabetes significantly modified VOC associated CAC depletion.

**Conclusion:** Low-level ambient exposure to VOCs is associated with CAC depletion, which could compromise endothelial repair and angiogenesis, and exacerbate CVD risk.

## 1. INTRODUCTION

Exposure to ambient air pollution has been widely recognized to be an important contributor to premature mortality and morbidity. The Global Burden of Disease (GBD) study estimates that in 2015, 4.2 million deaths could be attributed to ambient air pollutants, with an additional 2.8 million deaths due to household air pollution. ^1^ Exposure to fine particulate matter in the ambient air, defined as particle size < 2.5 μm in diameter (PM_2.5_), is associated with an increased risk for hypertension, ^2^ myocardial infarction, ^3^ type 2 diabetes, ^4, 5^ stroke, ^6^ and cardiovascular mortality. ^7^ The emphasis on PM_2.5_ and cardiovascular disease (CVD) has been motivated largely by the availability of historical monitoring data, however in an urban setting, over 90% of pollutant mass consists of gases or vaporous compounds, such as volatile organic compounds (VOCs). ^8^ In comparison with PM_2.5_, data on VOCs tend to exhibit higher temporal and spatial variability.

Most studies on VOC have focused on tobacco smoke ^9–11^ which represents a major source of human exposures. However, non-tobacco derived VOCs are ubiquitous in urban environments. High concentrations of VOCs such as alkanes, alkenes, aromatic hydrocarbons, and aldehydes, are emitted into the atmosphere from both anthropogenic and biogenic sources, and although biogenic VOCs are more abundant worldwide, in urban areas, anthropogenic VOCs are often higher. ^12^ Outdoor VOC exposures can originate from vehicle emissions, industrial facilities, landfills and hazardous waste sites, while indoor exposure can originate from diverse sources, such as paints, cleaning agents, personal care products, and solvents. ^13^ Our analysis of the secular trends of VOC exposures in the US suggests that even though there was a decline in the ambient air VOC levels, individual-level exposure increased from 2005-2014. ^14^ Hence, there is urgent need to assess the health impacts of VOC exposures.

VOC exposure have been associated with a wide range of acute and chronic health effects, including allergies, respiratory diseases, liver dysfunction, and cancer. ^15, 16^ Moreover, exposures to ambient levels of acrolein, benzene, and xylene have been associated with elevated CVD risk. ^17, 18^ In an intra-urban analysis of VOC gradients, higher levels of VOCs were found to be associated with increased cardiovascular and cancer mortality. ^16^ Moreover, longitudinal time-series studies have shown that ambient VOCs, such as alkenes and alkynes, are associated with increased heart failure emergency hospitalizations, ^19^ as well as overall cardiovascular hospitalizations, ^20^ and that occupational exposure to VOC such as styrene increases the relative risk of death from ischemic heart disease. ^21^

The biological plausibility that VOCs can induce cardiovascular injury and exacerbate CVD is extensively supported by a range of animal studies showing that exposures to benzene, acrolein, and formaldehyde, can lead to changes in blood pressure regulation, ^22–24^ depletion of circulating stem cells, ^25^ insulin resistance, ^26^ and platelet activation. ^27^ Exposure to these VOCs also induces systemic dyslipidemia, lipoprotein modification, ^28^ dilated cardiomyopathy; ^29^ and lead to the acceleration of atherogenesis ^30^ and exacerbation of myocardial ischemic injury. ^31^ Nonetheless, despite this evidence, little is known about the effects of VOCs on the mechanisms and processes that contribute to cardiovascular disease in humans. Previous work has shown that in humans, high levels of VOC exposures from sources such as cigarette smoke ^32, 33^ and diesel exhaust ^34^ are associated with endothelial dysfunction. We have previously found that exposure to acrolein and 1,3-butadiene are associated with lower reactive hyperemia index, a measure of endothelial function. ^13, 35^ The endothelium is a first line of defense of cardiovascular tissues against environmental toxins, and endothelial function is a sensitive reflection of vascular and systemic health. Therefore, to identify and understand the effects of VOC exposures and cardiovascular health, we measured circulating angiogenic cells (CAC), also referred to as endothelial progenitor cells (EPC). It is believed that these cell aid in the repair of the endothelium after injury, and are good biomarkers of vascular function. ^36, 37^ CAC levels are associated with several CVD risk factors, ^38^ and are predictive of both cardiovascular events and cardiovascular mortality. ^39^ Thus, using CACs as surrogate biomarkers of endothelial health, we examined their association with urinary levels of VOC metabolite levels in participants with low to severe CVD risk.

## 2. METHODS

### 2.1 Study Population and Design

In a cross-sectional study, adult study participants, aged 25-70 years, were recruited from the Louisville, KY area. Recruitment occurred in the summer months (May to September) in the years 2018 and 2019, to minimize seasonal outdoor VOC heterogeneity. Persons unwilling or unable to provide informed consent or with significant and/or severe comorbidities were excluded. These included subjects that had hepatitis, HIV/AIDS, active treatment for cancer, and active bleeding including wounds. Other exclusions included subjects with body weight less than 100 pounds, pregnant women, prisoners, and other vulnerable populations, those with conditions known to effect peripheral blood cell counts and bone marrow function.

At the time of enrollment non-fasting blood and spot urine were collected from patients who met the enrollment criteria and gave written consent. Demographic information, including age, sex, ethnicity, residential address, smoking status history and secondhand smoke exposure (verified by measurement of urinary cotinine concentrations), and BMI (calculated from self-reported height and weight) was obtained by using a questionnaire. Information on medication usage, blood pressure, and CVD history, including incidence of heart attack, heart failure, angina, hypertension, hypercholesterolemia, diabetes, obesity, stroke, and revascularization were also obtained through the questionnaire. For the current study, hypertension was defined as a previous diagnosis of hypertension or systolic blood pressure greater than 140 mmHg. Diabetes was defined as previous diagnosis of diabetes or HbA1C greater than 6.5. The study was approved by the University of Louisville Institutional Review Board (IRB 15.1260).

A total of 735 individuals met the inclusion criteria (Scheme 1).

**Scheme 1.**
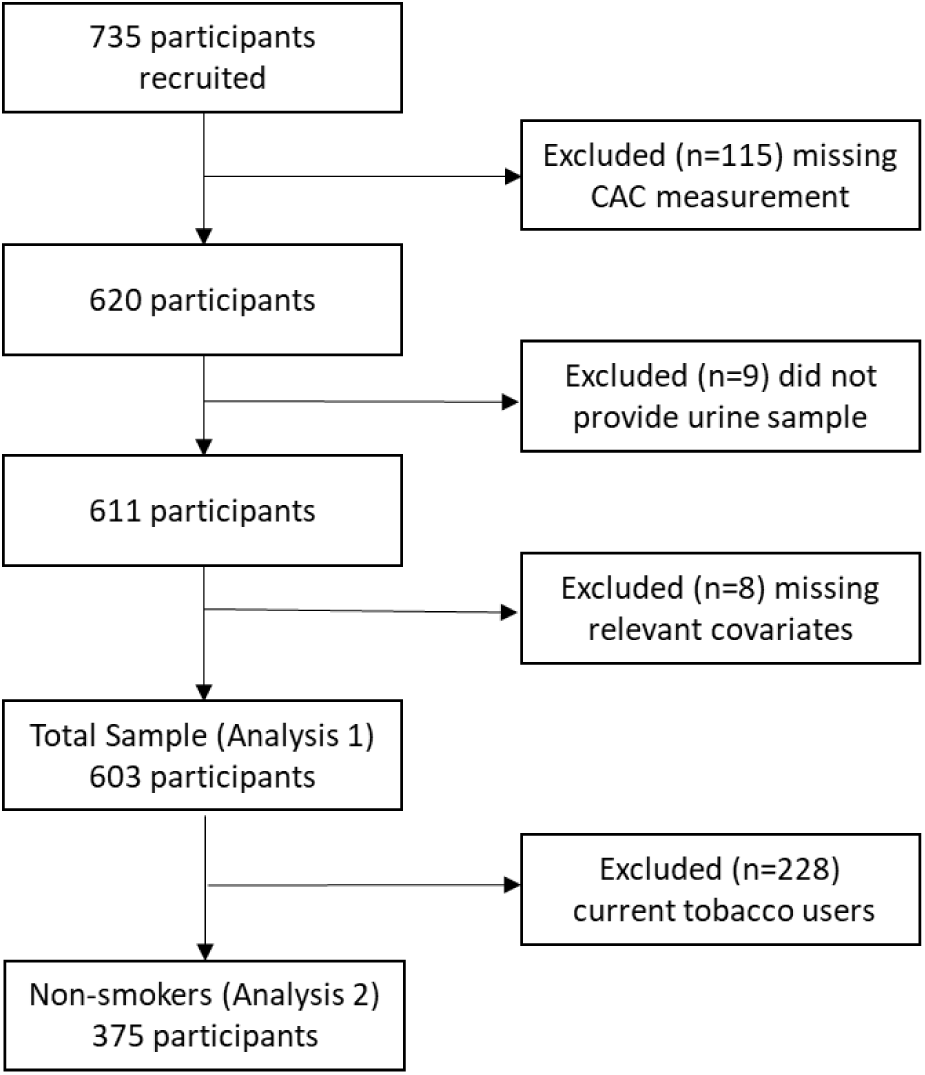
Recruitment Flow Chart.

Of those enrolled in the study, 115 individuals did not provide blood samples, or did not have circulating angiogenic cell measurements, 9 individuals did not provide urine samples, and 8 were missing other covariates. The final sample consisted of 603 participants with both urinary VOC metabolites and CAC measurements. For the non-smoker analysis, participants (n=228) with urinary cotinine value >40 µg/g creatinine were excluded, for a final sample size of n=375 participants.

### 2.2 Urinary Volatile Organic Compound Metabolites

Urinary metabolites of 12 VOCs (16 metabolites), and free forms of the nicotine metabolite cotinine were quantified by ultra-performance liquid chromatography-mass spectrometry (UPLC-MS/MS) as described before. ^9, 11, 40^ All VOC metabolites that were below the limit of detection (LOD) were imputed with the LOD value divided by the square root of 2. All urinary analytes were normalized to urinary creatinine levels, measured on a COBAS MIRA-plus analyzer (Roche, New Jersey) with Infinity Creatinine Reagent (Thermo Fisher Scientific, Massachusetts). A complete list of metabolites and parent compounds is provided in supplementary material (Table S1).

### 2.3 Circulating Angiogenic Cells

Fifteen different subtypes of circulating angiogenic cells were measured in the peripheral blood by flow cytometry - on fresh blood specimens as previously described. ^17, 41^ CACs were defined as CD14 negative cells and differentiated into subtypes based on expression of CD45, CD34, CD146 and AC133 surface markers.

### 2.4 Air Pollution and Meteorological Data

Data on ambient levels of PM_2.5_ (particulate matter with an aerodynamic diameter of ≤2.5 µm; µg/m^3^) and ozone (ppm) were obtained from three regional EPA-validated monitoring stations in the Louisville, KY region. We used the average of the three monitors for the day the study participants provided blood and urine samples.

Data on average daily temperature (°F) and relative humidity (%) were obtained from the National Oceanic and Atmospheric Administration’s (NOAA) National Centers for Environmental Information. Data was download for the Louisville International Airport monitoring station, which is in close proximity to the study participants’ residence, for the day the study participants provided blood and urine sample. This data is publicly available (https://www.ncdc.noaa.gov/cdo-web/datatools/selectlocation).

### 2.5 Statistical Analysis

Participant characteristics are expressed as mean ± standard deviation (SD) for continuous variables, and frequency (%) for categorical variables. Descriptive statistics were conducted for all VOC metabolites. Because CACs were positive, and heavily right-skewed, generalized linear models with the gamma distribution and log-link were used to examine the association between CACs and VOC metabolites (Figure 1). Models used natural log-transformed VOC metabolites and were adjusted based on prior literature showing an association with VOC exposure or CAC levels. ^41–47^ Model adjustments for full data set included: age, sex, race, BMI, diabetes status, hypertension status, daily PM_2.5_ levels, and smoking status. There are a total of 240 unique comparisons between CACs and VOC metabolites. To account for multiple testing, we applied the Benjamini-Hochberg procedure to control the false-discovery rate (FDR).

**Figure 1.**
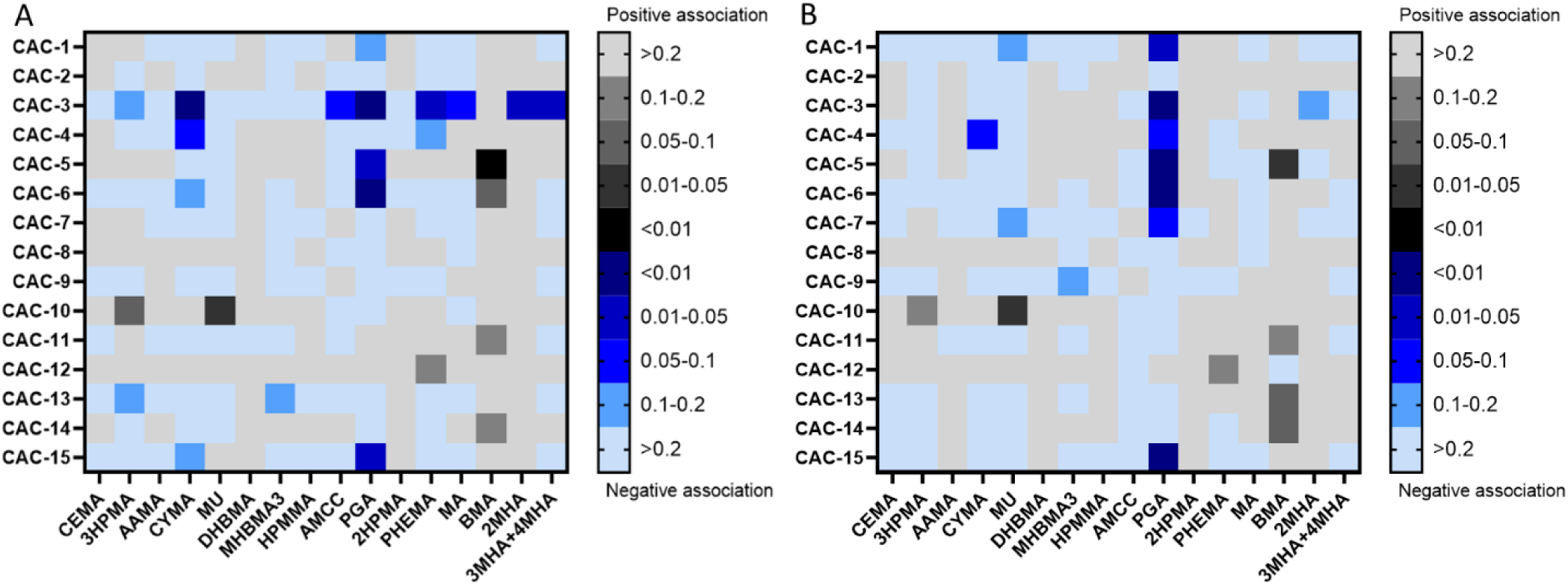
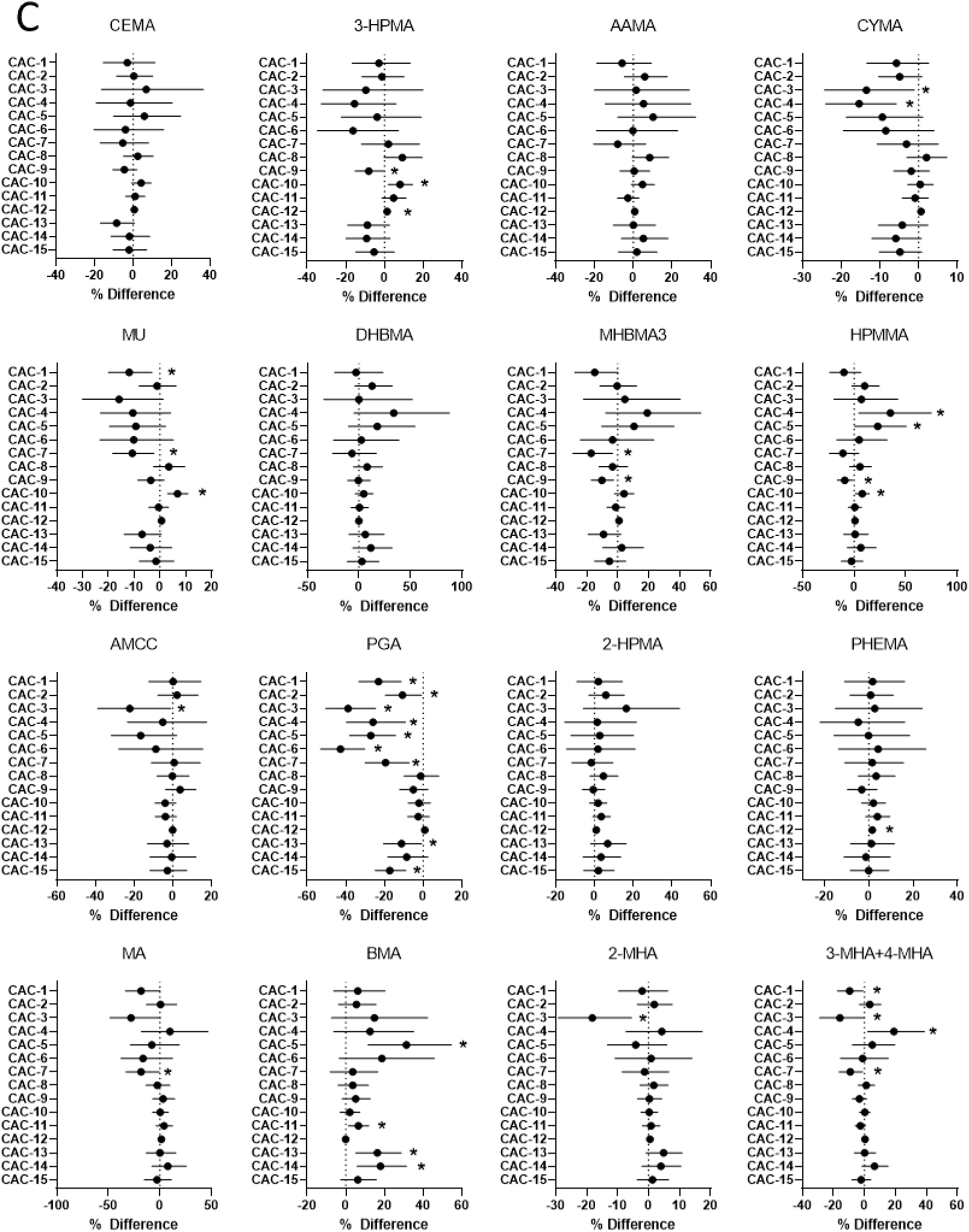
Association between the urinary metabolites of VOCs and circulating levels of CACs and heat map of p-values for pair-wise associations between the urinary metabolites of VOCs and circulating levels of CACs. (A) False Discovery rate adjusted p-values for total sample (n=603). (B) False Discovery rate adjusted p-values for non-smokers (n=375). Black grids represent positive associations and blue grids represent negative associations. The intensity of color represents p-values, with darker grids indicating smaller p-values. (C) The X axis represents % difference (± 95% confidence interval) per 2-fold increase in VOC metabolite for each CAC sub-population (CAC 1-15) in non-smokers (n=375). Generalized linear models were adjusted for age, sex, race, BMI, diabetes, hypertension, and daily PM_2.5_ levels. Models for the full sample were additionally adjusted for smoking status. Abbreviations: CEMA, N-Acetyl-S-(2-carboxyethyl)-L-cysteine; 3-HPMA, N-Acetyl-S-(3- hydroxypropyl)-L-cysteine; AAMA, N-Acetyl-S-(2-carbamoylethyl)-L-cysteine; CYMA, N-Acetyl-S- (2-cyanoethyl)-L-cysteine; MU, *trans, trans*-muconic acid; DHBMA, N-Acetyl-S-(3,4- dihydroxybutyl)-L-cysteine; MHBMA3, N-Acetyl-S-(4-hydroxy-2-buten-1-yl)-L-cysteine; HPMMA, N-Acetyl-S-(3-hydroxypropyl-1-methyl)-L-cysteine; AMCC, N-Acetyl-S-(N-methylcarbamoyl)-L- cysteine; PGA, Phenylglyoxylic acid ; 2HPMA, N-Acetyl-S-(2-hydroxypropyl)-L-cysteine; PHEMA, N-Acetyl-S-(1-phenyl-2-hydroxyethyl)-L-cysteine +; MA, Mandelic Acid; BMA, N-Acetyl-S- (benzyl)-L-cysteine; 2MHA, 2-Methylhippuric acid; 3MHA+4MHA, 3-Methylhippuric acid + 4- Methylhippuric acid. * Represents significant association.

#### Analysis of non-smokers

Models for non-smokers (urinary cotinine <40 µg/g creatinine) were adjusted for age, sex, race, BMI, diabetes status, hypertension status, and daily PM_2.5_ levels. Pearson correlations were performed across VOC metabolites and daily PM_2.5_, ozone, temperature, and relative humidity (Supplemental Figure 1). We examined potential non-linear relationships between VOC metabolites and CACs using restricted cubic splines, with tests for non-linearity using the likelihood ratio test. An environmental risk score (ERS) representing cumulative VOC exposure risk was calculated for each CAC using a modified version of the method developed by Park *et al.* ^48^ Briefly, (1) VOC metabolites were log transformed, (2) each log-transformed VOC metabolite was regressed against each CAC separately, while adjusting for covariates, (3) the environmental risk score for each participant was calculated as the sum of log-transformed VOC metabolite x (regression coefficient/standard error). If multiple VOC metabolites came from the same parent compound, only one was used in the creation of the ERS. The final set of VOC metabolites used to calculate the ERS consist of: 3HPMA, AAMA, CYMA, MU, MHBMA3, HPMMA, AMCC, PGA, 2HPMA, MA, BMA, and 3MHA+4MHA. Multi-pollutant models including all VOC metabolites that were independently associated with CACs were further developed to test whether single VOC effects were confounded by correlated VOCs (Pearson correlations provided in Supplemental Table 2). These models used log-transformed metabolite values, and the same adjustments as single pollutant models. Interactions between covariates and log VOC metabolites were tested in single pollutant models. We stratified analyses by participant characteristics to examine whether effect estimates differed by subgroups.

In addition to single-VOC models, we created models for multiple VOC exposures. To assess multiple VOC exposures as a mixture, we applied a Bayesian Kernel Machine Regression (BKMR) approach to flexibly model the relationship between the mixture of VOC metabolites and each of the 15 CACs. ^49^ The BKMR is a semi-parametric technique that models the exposure response relationship as a non-parametric kernel function of the mixture components, adjusting for covariates parametrically. Implementation of BKMR relied on a Gaussian kernel distribution, with flat priors. The Gaussian kernel flexibly captures underlying functional forms without specifying the shape of the exposure-response curve. This method can assess independent effects, estimate the overall mixture, and assess for interactions among mixture components. We used R software (version 4.0.2) and the bkmr package ^50^ to create posterior inclusion probabilities (PIPs) that quantify the relative importance of each exposure in the model, assess univariate and bivariate associations between mixture components and CACs, assess interactions among VOC metabolites, and estimate the effect of the overall mixture on individual CACs.

As a sensitivity analysis, we conducted generalized linear models additionally adjusting one at a time for temperature or relative humidity, while adjusting for the covariates as above. We then repeated the models adjusting for ozone in place of PM_2.5_, while adjusting one at time for temperature, then relative humidity. All statistical analyses were performed using SAS, version 9.4 (SAS Institute, Inc., Cary, North Carolina) and GraphPad Prism, version 8 (GraphPad Software, La Jolla, California).

## 3. RESULTS

The demographic characteristics of study participants are summarized in Table 1.

**Table 1.**
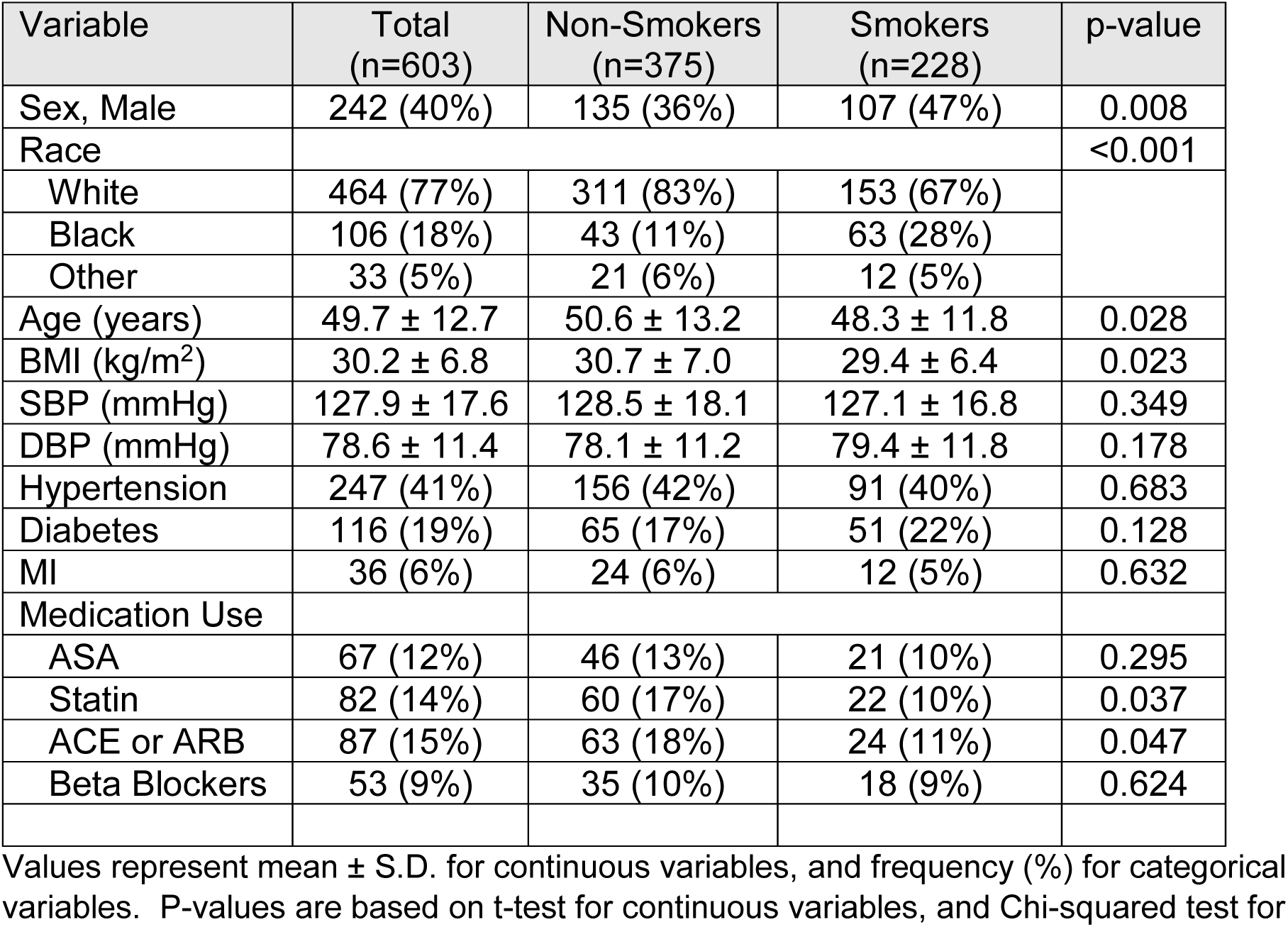
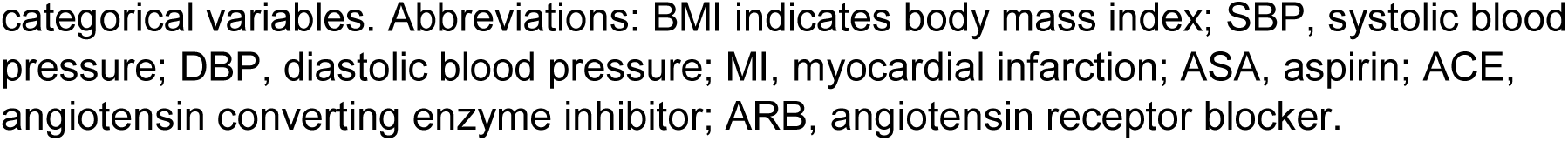
Participant Characteristics

| Variable | Total<br>(n=603) | Non-Smokers<br>(n=375) | Smokers<br>(n=228) | p-value |
| --- | --- | --- | --- | --- |
| Sex, Male | 242 (40%) | 135 (36%) | 107 (47%) | 0.008 |
| Race |  |  |  | <0.001 |
| White | 464 (77%) | 311 (83%) | 153 (67%) |  |
| Black | 106 (18%) | 43 (11%) | 63 (28%) |  |
| Other | 33 (5%) | 21 (6%) | 12 (5%) |  |
| Age (years) | 49.7 ± 12.7 | 50.6 ± 13.2 | 48.3 ± 11.8 | 0.028 |
| BMI (kg/m<sup>2</sup>) | 30.2 ± 6.8 | 30.7 ± 7.0 | 29.4 ± 6.4 | 0.023 |
| SBP (mmHg) | 127.9 ± 17.6 | 128.5 ± 18.1 | 127.1 ± 16.8 | 0.349 |
| DBP (mmHg) | 78.6 ± 11.4 | 78.1 ± 11.2 | 79.4 ± 11.8 | 0.178 |
| Hypertension | 247 (41%) | 156 (42%) | 91 (40%) | 0.683 |
| Diabetes | 116 (19%) | 65 (17%) | 51 (22%) | 0.128 |
| MI | 36 (6%) | 24 (6%) | 12 (5%) | 0.632 |
| Medication Use |  |  |  |  |
| ASA | 67 (12%) | 46 (13%) | 21 (10%) | 0.295 |
| Statin | 82 (14%) | 60 (17%) | 22 (10%) | 0.037 |
| ACE or ARB | 87 (15%) | 63 (18%) | 24 (11%) | 0.047 |
| Beta Blockers | 53 (9%) | 35 (10%) | 18 (9%) | 0.624 |

Values represent mean ± S.D. for continuous variables, and frequency (%) for categorical variables. P-values are based on t-test for continuous variables, and Chi-squared test for
categorical variables. Abbreviations: BMI indicates body mass index; SBP, systolic blood pressure; DBP, diastolic blood pressure; MI, myocardial infarction; ASA, aspirin; ACE, angiotensin converting enzyme inhibitor; ARB, angiotensin receptor blocker.

Compared with non-smokers, current smokers were more likely to be male (47% vs. 36%), Black (28% vs. 11%), were of younger age, and have lower BMI. Although cardiovascular risk factors (blood pressure, hypertension, diabetes) were similar between the two groups, non-smokers were more likely to use statins, and ACE or ARB medications. Within the 375 non-smoking participants of the study, the average age was 50.6 years. A majority were female (64%) and White (83%); 42% were hypertensive, and 17% diabetic. The urinary metabolites of VOCs measured in study participants are listed in Table 2, along with their parent VOCs.

**Table 2.**
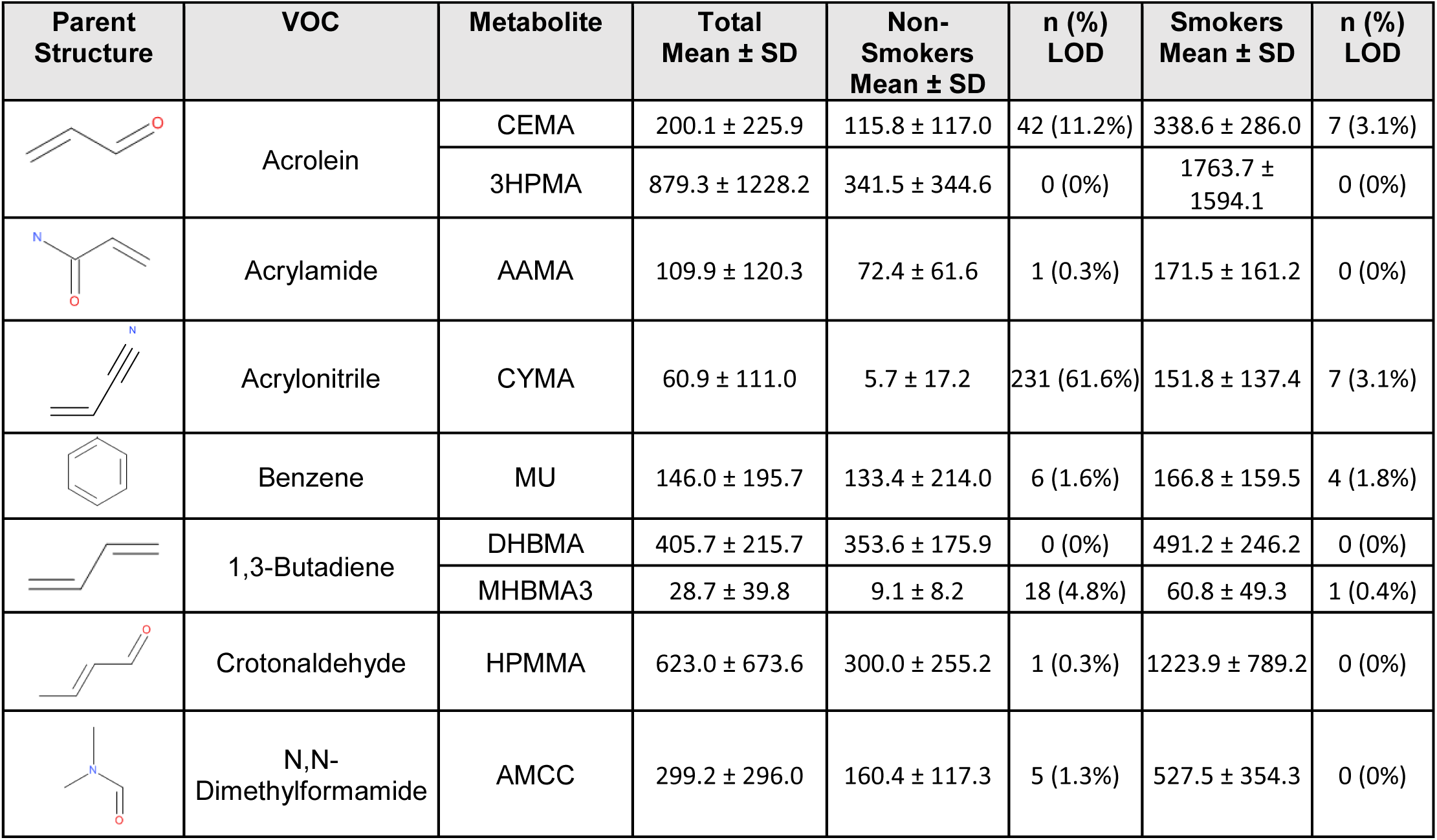
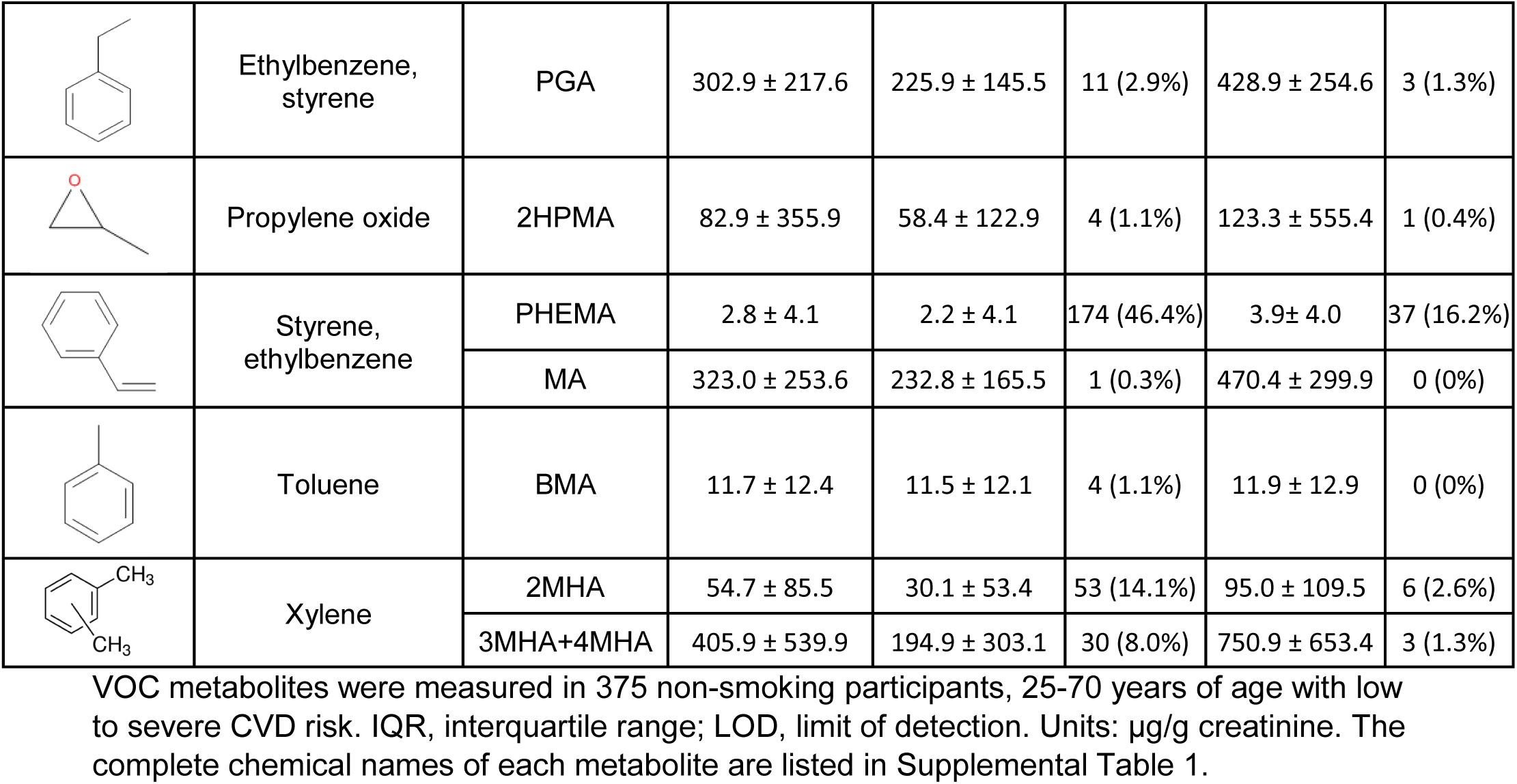
Urinary levels of the major metabolites of volatile organic compounds (VOCs) in study participants.

| Parent Structure | VOC | Metabolite | Total Mean ± SD | Non-Smokers Mean ± SD | n (%) LOD | Smokers Mean ± SD | n (%) LOD |
| --- | --- | --- | --- | --- | --- | --- | --- |
| | Acrolein | CEMA | 200.1 ± 225.9 | 115.8 ± 117.0 | 42 (11.2%) | 338.6 ± 286.0 | 7 (3.1%) |
| | | 3HPMA | 879.3 ± 1228.2 | 341.5 ± 344.6 | 0 (0%) | 1763.7 ± 1594.1 | 0 (0%) |
| | Acrylamide | AAMA | 109.9 ± 120.3 | 72.4 ± 61.6 | 1 (0.3%) | 171.5 ± 161.2 | 0 (0%) |
| | Acrylonitrile | CYMA | 60.9 ± 111.0 | 5.7 ± 17.2 | 231 (61.6%) | 151.8 ± 137.4 | 7 (3.1%) |
| | Benzene | MU | 146.0 ± 195.7 | 133.4 ± 214.0 | 6 (1.6%) | 166.8 ± 159.5 | 4 (1.8%) |
| | 1,3-Butadiene | DHBMA | 405.7 ± 215.7 | 353.6 ± 175.9 | 0 (0%) | 491.2 ± 246.2 | 0 (0%) |
| | | MHBMA3 | 28.7 ± 39.8 | 9.1 ± 8.2 | 18 (4.8%) | 60.8 ± 49.3 | 1 (0.4%) |
| | Crotonaldehyde | HPMMA | 623.0 ± 673.6 | 300.0 ± 255.2 | 1 (0.3%) | 1223.9 ± 789.2 | 0 (0%) |
| | N,N-Dimethylformamide | AMCC | 299.2 ± 296.0 | 160.4 ± 117.3 | 5 (1.3%) | 527.5 ± 354.3 | 0 (0%) |
|  | Ethylbenzene,<br>styrene | PGA | 302.9 ± 217.6 | 225.9 ± 145.5 | 11 (2.9%) | 428.9 ± 254.6 | 3 (1.3%) |
|  | Propylene oxide | 2HPMA | 82.9 ± 355.9 | 58.4 ± 122.9 | 4 (1.1%) | 123.3 ± 555.4 | 1 (0.4%) |
|  | Styrene,<br>ethylbenzene | PHEMA | 2.8 ± 4.1 | 2.2 ± 4.1 | 174 (46.4%) | 3.9 ± 4.0 | 37 (16.2%) |
|  |  | MA | 323.0 ± 253.6 | 232.8 ± 165.5 | 1 (0.3%) | 470.4 ± 299.9 | 0 (0%) |
|  | Toluene | BMA | 11.7 ± 12.4 | 11.5 ± 12.1 | 4 (1.1%) | 11.9 ± 12.9 | 0 (0%) |
|  | Xylene | 2MHA | 54.7 ± 85.5 | 30.1 ± 53.4 | 53 (14.1%) | 95.0 ± 109.5 | 6 (2.6%) |
|  |  | 3MHA+4MHA | 405.9 ± 539.9 | 194.9 ± 303.1 | 30 (8.0%) | 750.9 ± 653.4 | 3 (1.3%) |

VOC metabolites had a high degree of variability in both smokers and non-smokers. Participants who were defined as current smokers consistently showed higher average VOC metabolite levels than non-smokers. Although significant levels of most VOC metabolites could be detected in non-smokers, in many (>20%) samples the level of acrylonitrile (CYMA), styrene/ethylbenzene (PHEMA) metabolites were below the limit of detection (LOD).

In single-VOC analysis, the levels of several urinary VOC metabolites were associated with the levels of CACs measured in peripheral blood (Figure 1). In the full data set, PGA was significantly associated with CAC-3, 5, 6, and 15, MU was significantly associated with CAC-10, and BMA was associated with CAC-5. Additionally, we found that CAC-3 was associated with CYMA, PHEMA, 2-MHA, and 3-MHA+4-MHA, suggesting that CAC-3 may be a sensitive marker to tobacco derived VOCs.

### Analysis of non-smokers

In our analysis of non-smokers (Figure 1B, 1C), we found similar associations as in our analysis of the total sample. In non-smokers, some of the most pronounced effects were observed with PGA – a metabolite that could be derived from either ethylbenzene or styrene - which showed a significant inverse association with CACs-1,2,3,4,5,6, and 7, as well as CAC-13 and CAC-15. Several other VOC metabolites showed similar, but variable associations. The percent difference ranged from -10.7% (95% CI: -19.6%, - 0.8%) to -42.8% (95% CI: -53.1%, -30.1%). Many VOC metabolites were inversely associated with CAC levels, including metabolites of xylene (3-MHA+4-MHA), acrylonitrile (CYMA), and 1,3-butadiene (MHBMA3). For benzene (MU), we observed a significant inverse association with CAC-1 (-11.9%; 95% CI: -20.0%, -3.0%), and its subpopulation CAC-7 (-10.7%; 95% CI: -18.3%, -2.3%), along with a positive association with CAC-10. Positive associations were also observed with toluene (BMA) and CAC-5, 11, 13, and 14, which share AC133^+^ marker. After correcting for multiple testing, PGA showed statistically significant associations with CAC-1, 3, 5, 6, and 15 (Figure 1B). Further, MU remained significantly associated with CAC-10 and BMA remained significantly associated with CAC-5.

In our sensitivity analysis of non-smokers, additionally adjusting temperature, relative humidity, and ambient ozone did not substantially alter our associations (Supplemental Figure 2). Overall, these observations suggest that exposure to several VOCs is generally associated with lower levels of several CAC subpopulations in peripheral blood, particularly the early progenitor CACs.

Because we found similar associations between CACs and VOCs in the total sample, and in non-smokers, we focused the rest of our analysis on the subgroup of non-smokers, to determine how low levels of VOCs are associated with endothelial damage. In non-smokers, there was minimal correlation between VOC metabolites and average daily ambient PM_2.5_, ozone, temperature, and relative humidity (Supplemental Figure 1). The levels of urinary VOC metabolites showed a strong dependence on demographics. A comparison of urinary metabolites showed that females had higher levels of the metabolites of N,N’-DMF (AMCC), ethylbenzene/styrene (phenylglyoxylic acid, PGA and mandelic acid, MA) and toluene (BMA) than males. Likewise, in comparison with Blacks, Whites had higher levels of metabolites of benzene (*trans, trans*-muconic acid, MU), N,N’- DMF (N-Acetyl-S-(N-methylcarbamoyl)-L-cysteine, AMCC) and styrene/ethylbenzene (MA). The metabolites of several VOCs – 1,3, butadiene (N-Acetyl-S-(3,4-dihydroxybutyl)-L-cysteine, DHBMA), crotonaldehyde (N-Acetyl-S-(3-hydroxypropyl-1-methyl)-L-cysteine, HPMMA), N,N’-DMF (AMCC), styrene/ethylbenzene (MA), toluene (BMA), and xylene (3MHA+4MHA) were also higher in those >55 years of age. Urinary levels of the metabolite of butadiene (MHBMA3) and toluene (N-Acetyl-S-(benzyl)-L-cysteine, BMA and, 2-Methylhippuric acid, 2MHA) levels were higher in those with a BMI of <30 (Supplemental Table 2), reflecting different levels of exposure.

To determine the extent to which total VOC exposure can affect CAC levels, and which cell populations contribute to the directionality of the associations (found in Figure 1) we created an Environmental Risk Score (ERS) for each cell population (Figure 2).

**Figure 2.**
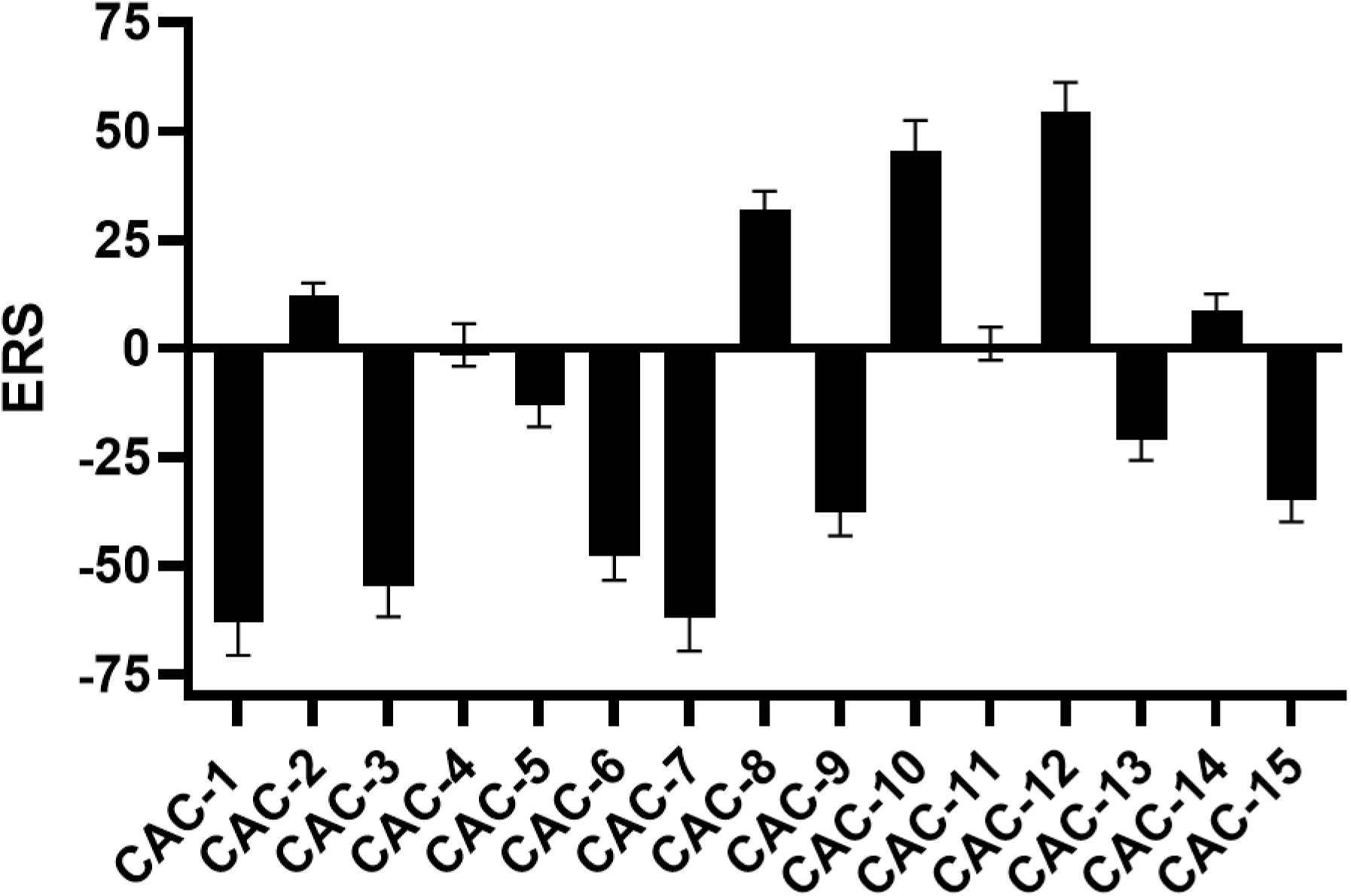
Association between Environmental Risk Scores and Circulating Angiogenic Cells in non-smokers. Environmental Risk score was calculated for each cell population as follows: 1. The urinary levels of VOC metabolites were log transformed. 2. Each log transformed VOC metabolite level was regressed against the levels of the cells separately, while adjusting for covariates. 3. ERS was calculated as the sum of the log transformed VOC metabolite levels x (regression coefficient/standard error). Mean and standard deviation of the ERS was calculated for each CAC. Values are mean ± SD of environmental risk score for each cell population.

We found a strong negative risk score for CAC-1 (-62.9), CAC-3 (-54.6), and CAC-7 (-62.1), which are a related group of cells with common markers CD45^dim^/CD146^+^/CD34^+^. To a lesser extent, we also found negative risk scores for CAC-6 (-47.6), and CAC-9 (-37.7), which are CD34^+^ cells. In comparison, we found positive risk scores for CD45^+^ cells (CAC-12), along with subgroups of CD45^+^ cells: CAC-2, CAC-8, and CAC-14. Moreover, we also found a positive score for CAC-10, which is CD146^+^. This indicates that the levels of CD45^+^ cells were positively associated with the urinary metabolites of VOC metabolites, whereas CD45^dim^ and CD34^+^ cells were negatively associated with the metabolite levels.

We used restricted cubic splines to determine whether the relationship between VOC metabolites and CACs was non-linear (Figure 3).

**Figure 3.**
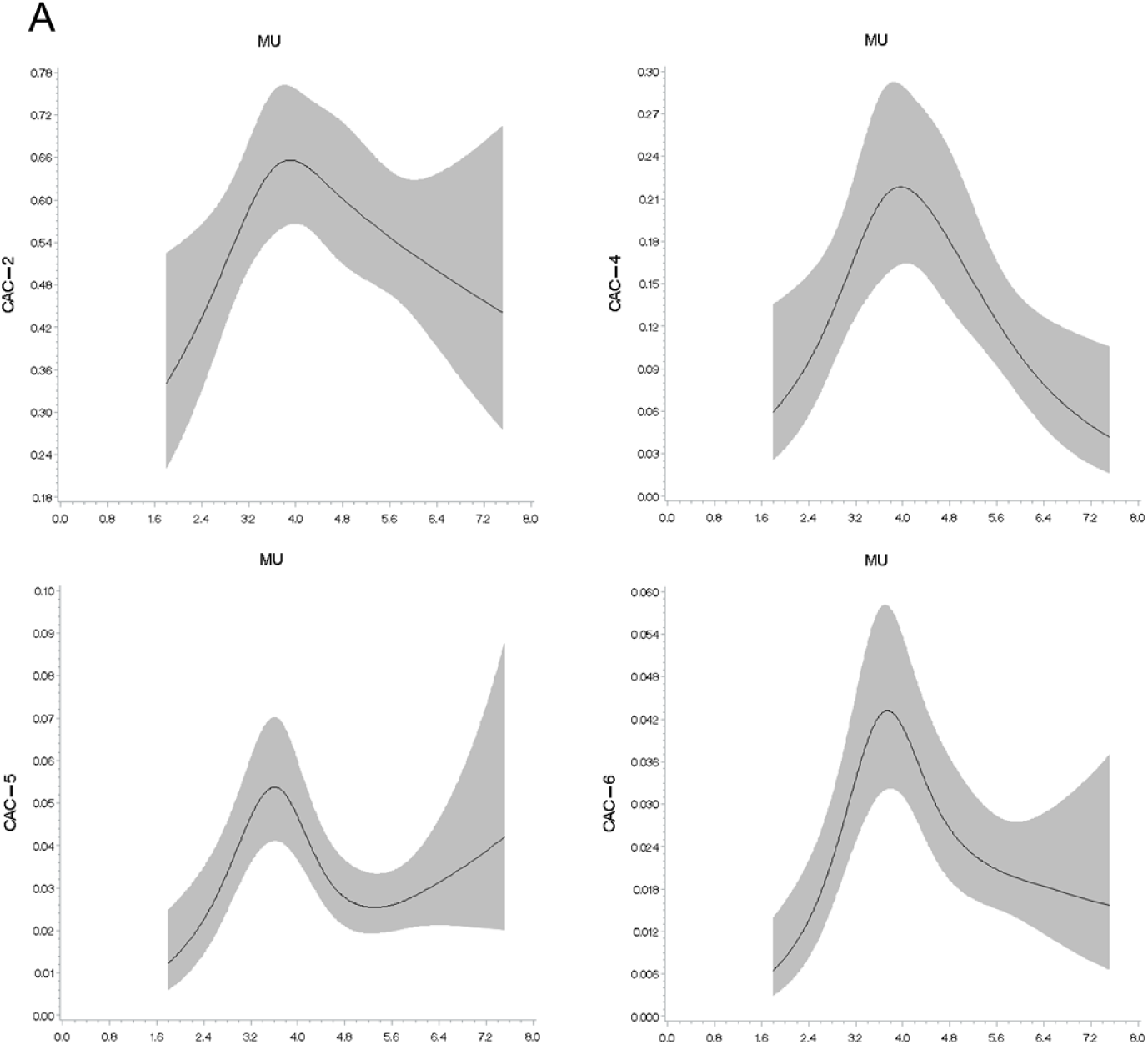
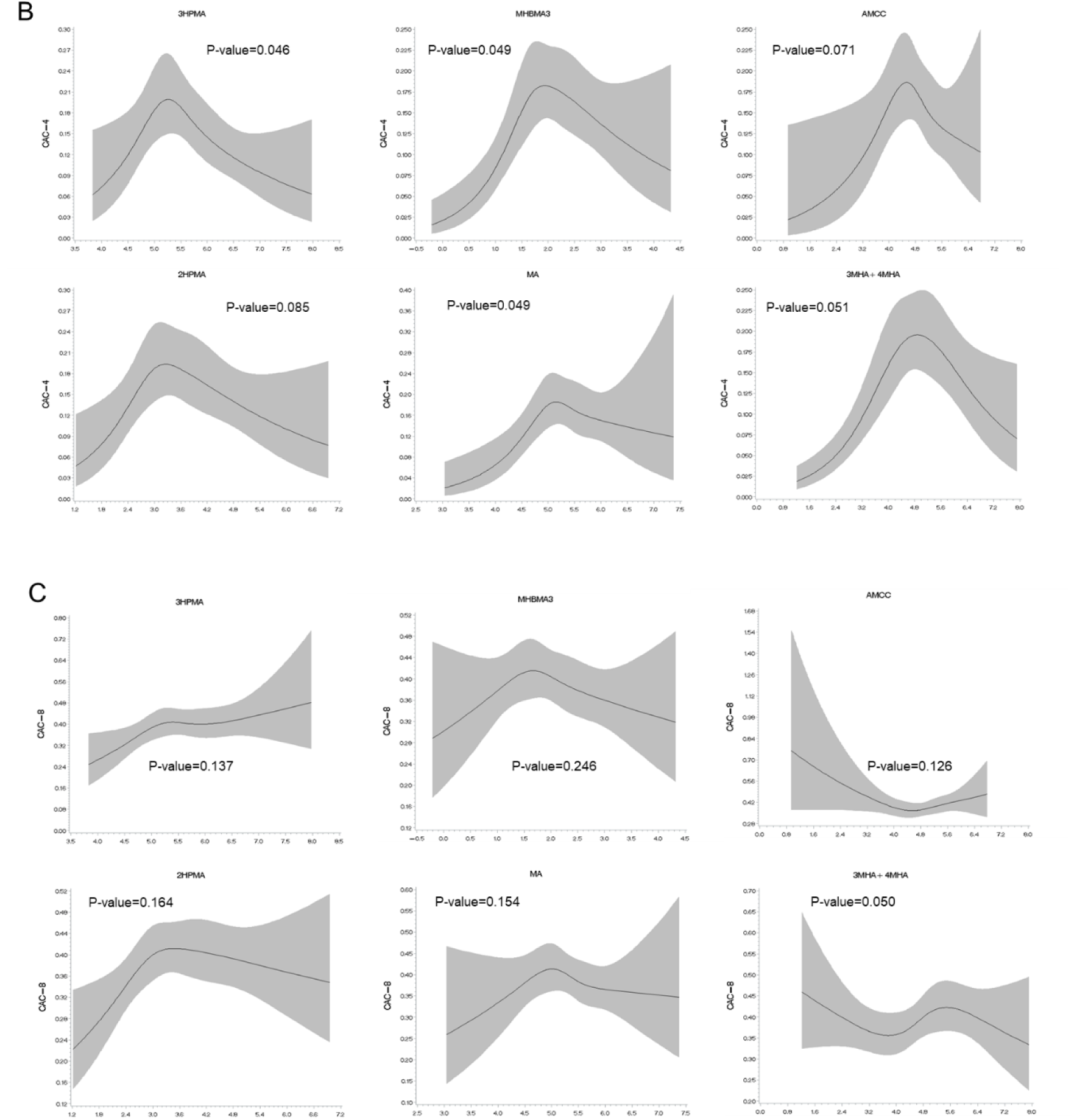
Exposure-response relationships between selected CACs and VOCs in non-smokers. Non-linear relationships between MU metabolites and CACs (**A**); VOCs and CAC-4 (**B**); and VOCs and CAC-8 (**C**) were examined using restricted cubic splines. Tests for non-linearity used the likelihood ratio test. Models are adjusted for age, sex, race, BMI, diabetes, hypertension, beta-blocker use, and daily PM_2.5_ levels. P-value represents likelihood ratio test for non-linearity.

We observed non-linear relationships between the circulating levels of several cell populations and the metabolite of benzene (MU). In general, low levels of MU was associated with higher levels of circulating cells, whereas higher MU levels were associated with lower cell levels. These non-linear relationships are presented in Figure 3A, which shows a near significant (p<0.1) non-linear relationships with CD146+ cell populations (CAC-2, 4, 5, and 6), and with AC133+ cell population – CAC-4 and CAC-5.

To determine whether AC133^+^ cells contribute to the non-linear relationships, we examined the association of subgroups of CD146^+^ cells, CAC-4 (CD45^+^/CD146^+^/AC133^+^) and CAC-8 (CD45^+^/CD146^+^/AC133^-^) with VOC metabolites. We found near significant (p<0.1) non-linear relationships between CAC-4 and metabolites of acrolein, 1,3- butadiene, N, N-dimethylformamide, propylene oxide, ethylbenzene/styrene, and xylene. These non-linear relationships were not observed with CAC-8, with the exception of xylene (3-MHA+4-MHA), suggesting that early progenitor endothelial cells (CD146^+^/AC133^+^) are recruited at low exposure levels, but are depleted at high VOC exposures.

Given that co-exposure to VOCs is common, we used multi-VOC models to assess whether the significant VOC effects found in single-VOC models were confounded by co-exposures to other VOCs (Figure 4).

**Figure 4.**
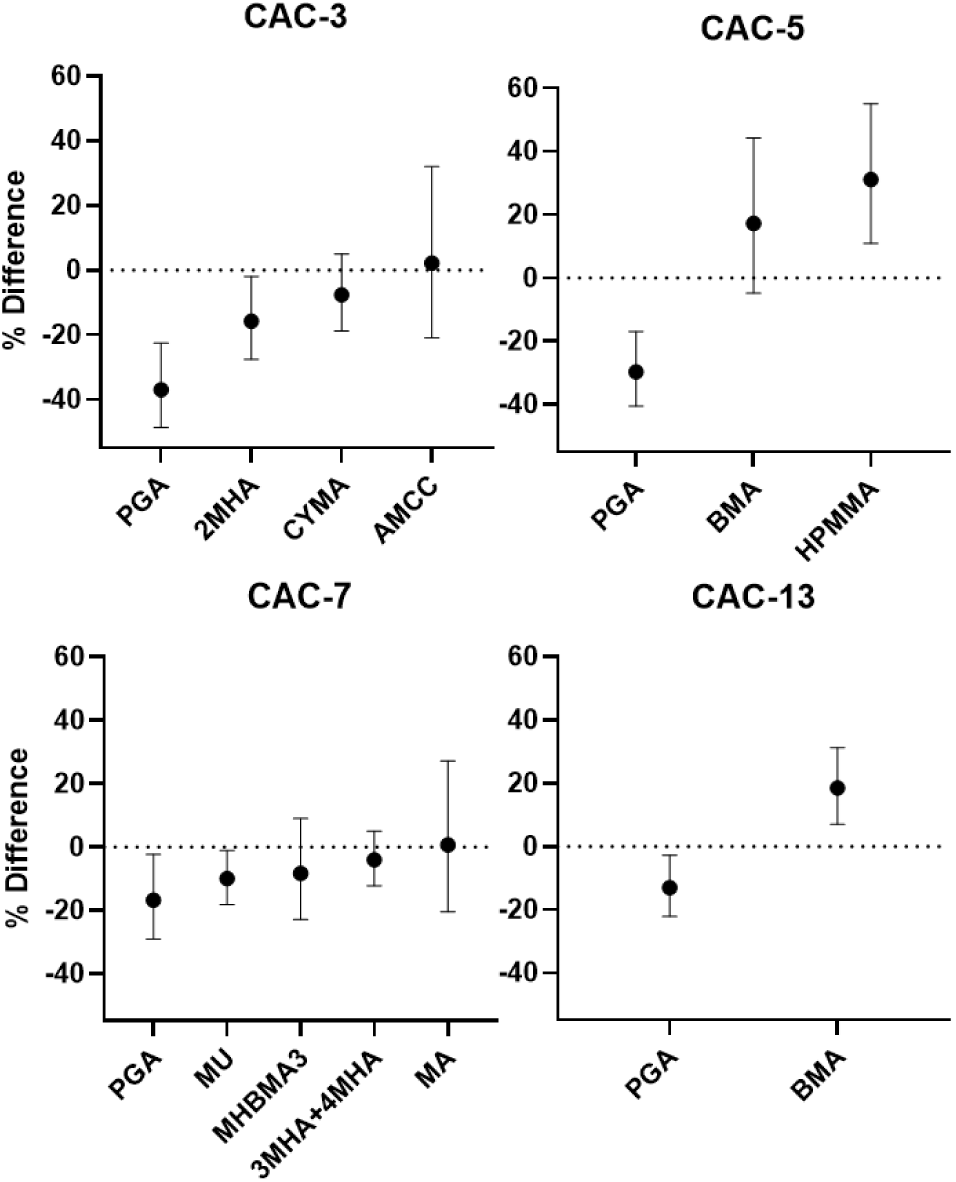
Association between circulating angiogenic cells and multiple VOCs in the same model (non-smokers). Values represent % difference per 2-fold increase in VOC metabolite. VOC metabolites are ordered by strength of their regression coefficients. Only significant metabolites from one-pollutant models were entered in the model. Models adjusted for age, sex, race, BMI, diabetes, hypertension, and daily PM_2.5_ levels.

For this analysis, we focused on rare cell populations with negative ERS scores CAC-3, CAC-5, CAC-7, and CAC-13 (Figure 2). We observed that the metabolite of ethylbenzene/styrene (PGA) remained negatively associated with CAC-3 (-37.0% per 2- fold increase; 95% CI: -48.7%, -22.6%), CAC-5 (-29.8%; 95% CI: -40.6%, -17.0%), CAC-7 (-16.8%; 95% CI: -29.1%, -2.4%), and CAC-13 (-13.1%; 95% CI: -22.2%, -2.8%), suggesting that the associations between PGA and CACs are independent of other VOC metabolite levels. We also found that the metabolites of xylene (2MHA) remained negatively associated with CAC-3, crotonaldehyde (HPMMA) remained positively associated with CAC-5, benzene (MU) remained negatively associated with CAC-7, and toluene (BMA) remained positively associated with CAC-13.

In our Bayesian kernel machine regression analysis, important VOC metabolites contributing to changes in CACs are indicated by high posterior inclusion probabilities (Table 3).

**Table 3.**
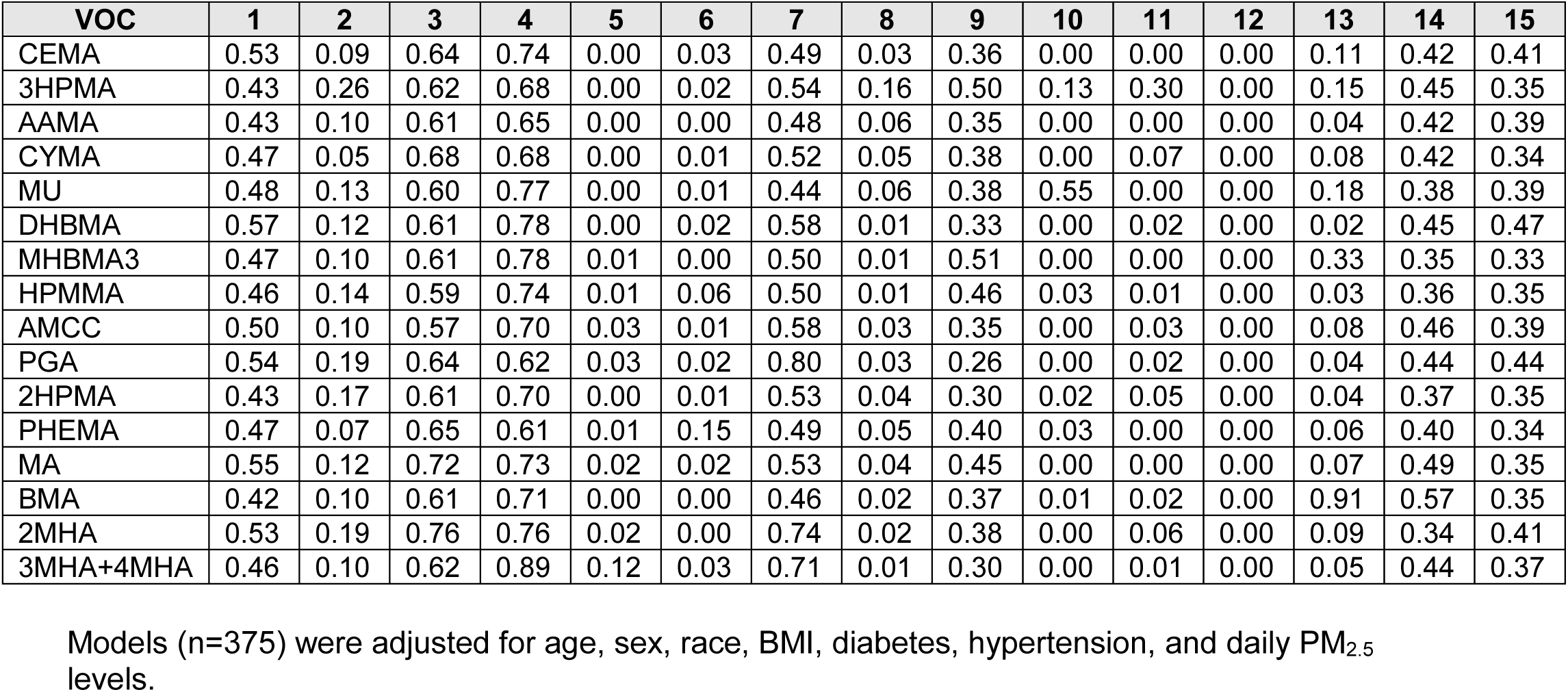
Posterior inclusion probabilities of individual VOC metabolites in the BKMR model for 15 CACs.

The directionalities of single VOC associations in our mixture models were generally consistent with the observed directionalities in our single VOC generalized linear models. For example, in mixture models, individuals with greater PGA levels had lower numbers of CACs, while individuals with greater BMA and HPMMA levels had higher numbers of CACs. As evident in Figure 5, we also found that the mixture of VOCs had a nonlinear relationship with CAC-4, similar to the nonlinear associations observed in our single VOC models. We also found that greater exposure to the VOC mixture was associated with higher CAC-10 levels (Figure 5).

**Figure 5.**
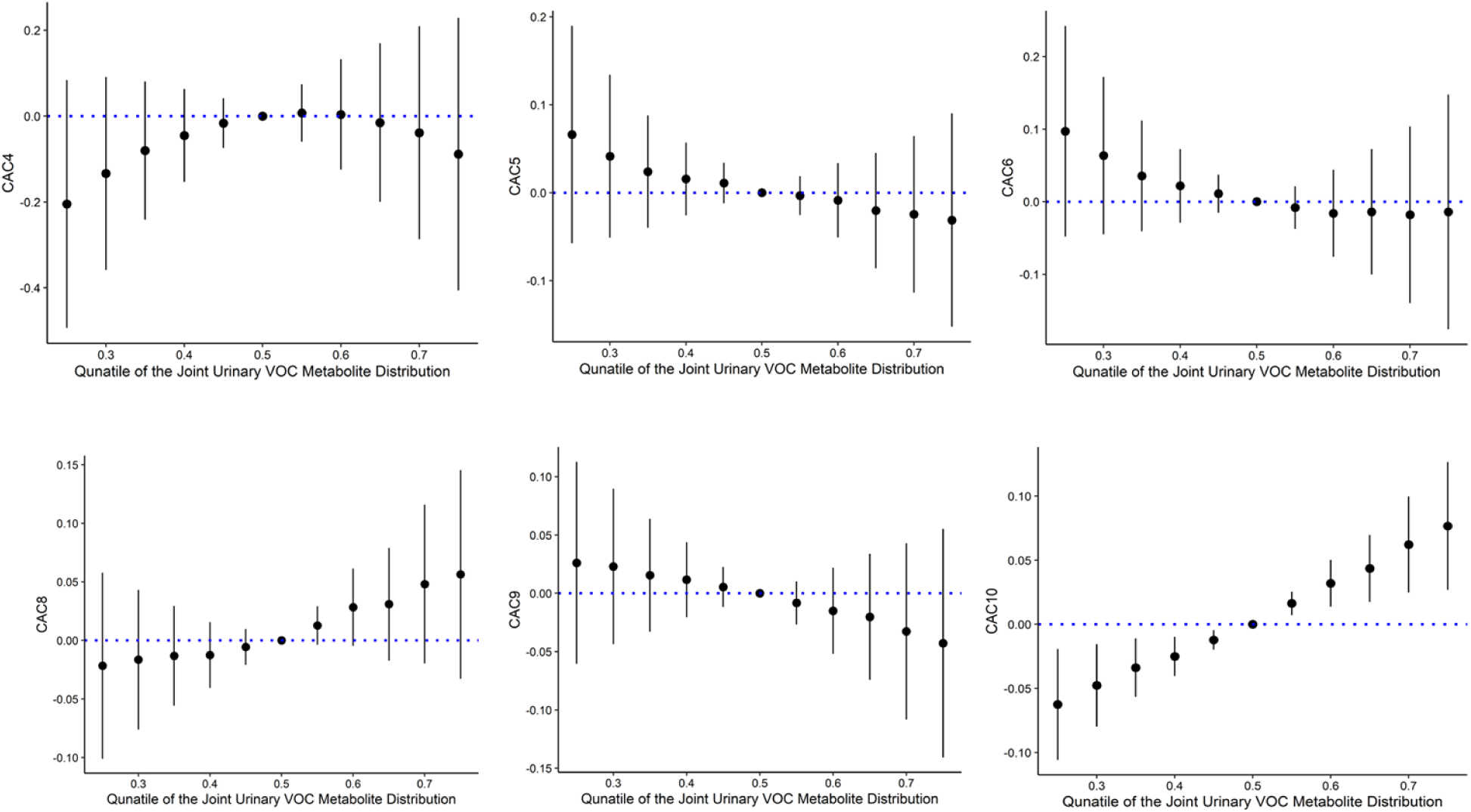
Overall association of the VOC metabolite mixture (and 95% credible intervals) with selected CACs, when all exposures are set to a given percentile relative to median. Models (non-smokers; n=375) were adjusted for age, sex, race, BMI, diabetes, hypertension, and daily PM_2.5_ levels.

The effect of the VOC mixture on CAC-10 was largely driven by benzene (MU), with a posterior inclusion probability of 0.55, where all other VOC metabolites were less than 0.15. We found minimal evidence of interactions between pairs of VOC metabolites in our mixture analysis (data not shown).

Because the metabolites of benzene, 1,3-butadiene, N,N-dimethylformamide, and ethylbenzene/styrene consistently showed strong associations with CACs, we investigated whether these associations were modified by the demographic characteristics of the participants or their co-morbidity profile. In our subgroup analysis, we found a common theme in many of the models, in which participants who were male, White, normotensive, and recruited on low PM_2.5_ days showed stronger inverse relationships between VOCs and CAC levels (Figure 6).

**Figure 6.**
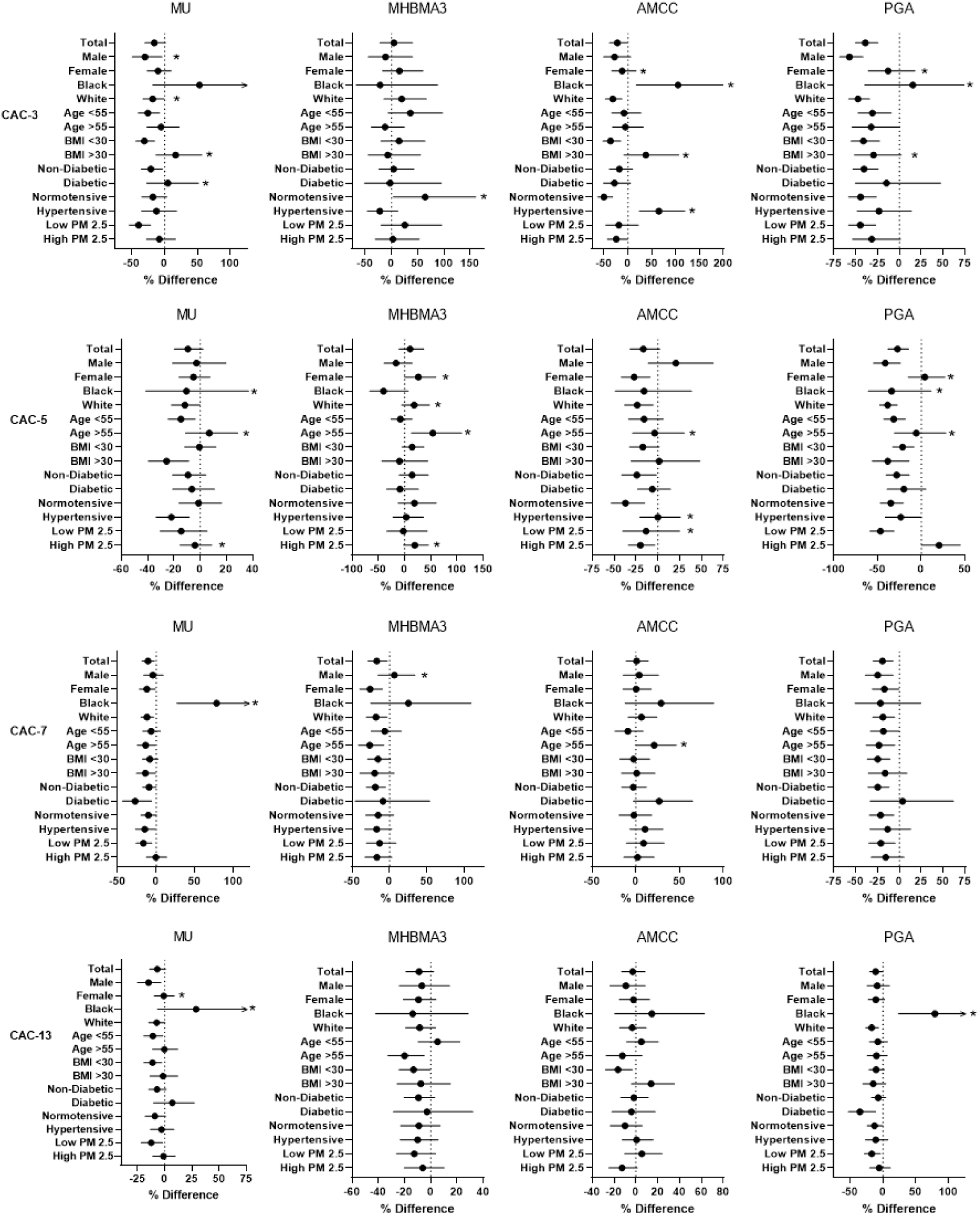
Subgroup analysis of the relationship between different CAC population with the urinary metabolites of benzene (MU), 1,3-butadiene (MHBMA3), ethylbenzene/styrene (PGA), and N,N-Dimethylformamide (AMCC) in non-smokers. Generalized linear models were adjusted for covariates -age, sex, race, BMI, diabetes, hypertension, and daily PM levels, unless the covariates were used for stratification. *Significant interaction between subgroups in the full model.

For example, for CAC-13, we found a negative association between ethylbenzene/styrene metabolite (PGA) in White participants of -17.2% per 2-fold increase in CAC levels (95% CI: -26.4%, -7.0%), whereas in Black participants, we found a positive association of 79.1% (95% CI: 23.7%, 159.3%). We observed a similar relationship for the metabolite of benzene (MU), when stratified by race; however, this may be due to lower levels of MU in urine samples from Black participants (62.2 μg/g creatinine) than White participants (146.0 μg/g creatinine), as presented in *Supplemental Table 2*.

We also found some evidence that age and BMI modify the relationships between VOCs and CAC levels, although the direction was not consistent in all models. Interestingly, in several of our models, we found stronger inverse associations between VOC metabolite and CACs in normotensives than hypertensives, and to a lesser extent, non-diabetics than diabetics, - conditions that are known to affect cell migration. For example, for CAC-5, we found a negative association with ethylbenzene/styrene metabolite (PGA) in non-diabetics of -28.2% per 2-fold increase, and with N,N-dimethylformamide (AMCC) in non-diabetics of -24.0% per 2-fold increase, while the relationships in diabetics were not significant. To determine whether these results were due to differential levels of VOC exposure between subgroups, we calculated average metabolite levels for each subgroup *Supplemental Table 2*. We found that levels of MU (benzene) and PGA (ethylbenzene and styrene) were higher in diabetics than non-diabetes and were higher in hypertensive than normotensive individuals, although normotensive had higher levels of 2HPMA (propylene oxide).

## 4. DISCUSSION

The major findings of this study are that the urinary metabolites of several common VOCs, such as ethylbenzene/styrene, benzene, xylene, and 1,3-butadiene were negatively associated with circulating angiogenic cells. We found similar associations in non-smokers compared with the total sample containing smokers. For some VOCs, such as benzene, we observed that low levels of their metabolites were positively associated with an increase in CAC levels, whereas at high levels of exposure were associated with a significant decrease in CAC levels. Our environmental risk score identified that CAC-1, CAC-3, and CAC-7, with common cell markers CD45^dim^/CD146^+^/CD34^+^, were the most sensitive to total VOC exposure. Our mixture analysis revealed that the total VOC metabolite mixture was positively associated with CD146^+^ cells. We also found sex, race, age, and hypertension to be effect modifiers of the some of the relationships between the levels of VOC metabolites and CACs. Taken together, these findings suggest that exposure to VOCs could lead to deficits in endothelial repair capacity in a dose-dependent manner, which vary with sex, race, and comorbidities. Given that low CAC levels are predictive of future cardiovascular events and death ^39^, these results support the notion that VOC exposure is associated with significant vascular injury independent of tobacco use, and therefore it may be an important environmental risk factor for CVD.

The potential cardiovascular effects of VOCs derived from tobacco smoke have been extensively studied. ^9–11^ We found similar associations between VOCs and CACs in our total sample containing smokers, compared with our subgroup of non-smokers. This suggests that the effects of VOC exposure on CACs is independent of tobaccos smoke exposure and is still present at environmentally low levels. We additionally found that several VOC metabolites were associated with CAC-3 in the total sample analysis. We have previously shown that low levels of CAC-3 is associated with higher prevalence of diabetes, higher HbA1c levels, and impaired vascular function.^37^ This could be due to the higher levels of several VOC metabolites in the total sample, such as CYMA (acrylonitrile), and PHEMA (ethylbenzene/styrene), which are typically difficult to detect in non-smokers. In contrast, the cardiovascular effects of VOCs not derived from tobacco smoke have received scant attention. Nonetheless, in addition to tobacco smoke, automobile emissions, industrial facilities, hazardous waste sites, and household products are major sources of VOC emissions, and therefore, VOC exposures are common and frequent. ^13^ In our study, we found significant levels of the VOC metabolites in the urine of non-smokers, indicating particularly high levels of exposure to acrolein, crotonaldehyde, and 1,3-butadiene, and acrylamide, as well as the BTEXS (benzene, toluene, ethylbenzene, xylene and styrene) group of VOCs, suggesting a contribution of both indoor and outdoor exposure. Specifically, we found that the CAC levels were particularly sensitive to metabolites of BTEXS compounds. We observed significant associations between CACs and urinary metabolite of benzene (3 CACs), toluene (4 CACs), ethylbenzene/styrene (9 CACs), and xylene (4 CACs).

Although exposure to BTEXS is common, little is known about their cardiovascular effects. A study from Hong Kong found that short term elevations in atmospheric levels of benzene and TEX (toluene, ethylbenzene, and xylene) was associated with increases in circulatory disease mortality by 5.8% and 3.5%, ^51^ although in a study from Toronto, no intra-urban variations in VOC levels were found be associated with cardiovascular disease mortality. ^16^ Other BTEXS such as toluene, styrene, ethylbenzene, and xylenes have been also linked to a higher prevalence of CVD, ^52^ and exposure to VOCs such as benzene, ethylbenzene, and styrene have also been found to be associated with higher odds of developing metabolic syndrome. ^53^ Along with these data, the results of our study, support the notion that exposure to VOC could have adverse cardiovascular consequences, potentially due to low-level vascular injury reflected by changes in CAC levels.

We found inverse associations between metabolites of acrylonitrile and 1, 3- butadiene and CACs. Although the cardiovascular effects of acrylonitrile are less well studied, exposure to 1,3-butadiene has been found to accelerate the formation of atherosclerotic lesions in animal models, and polymorphisms in phase II enzymes that metabolize 1,3-butadiene are associated with the development of atherosclerosis in humans. ^54^ Hence, our observation that urinary metabolites of 1,3-butadiene are associated with CAC levels suggest that the cardiovascular risk of exposure to this VOC may be in part due to depletion of early, progenitor cells, which aid in vascular repair and thereby can prevent the progression of atherosclerotic lesions. Therefore, our observations linking exposure to VOCs such as 1,3-butadiene to concurrent changes in CACs, not only provides a new biological basis for the cardiovascular effects of VOCs, independent of concurrent exposure to PM_2.5_, but also provides new insights into their mechanism of toxicity.

In real world scenario, the urban air contains many different VOCs and therefore, we conducted multi-VOC models controlling for all significantly associated metabolites from the single VOC models. In particular, we found that the urinary metabolite PGA remained significantly associated with the levels of CD45^dim^/CD146^+^/CD34^+^ cells (CAC-3, CAC-7), AC133^+^/CD146^+^ cells (CAC-5), and AC133^+^/CD34^+^ cells (CAC-13). PGA is oxidized from mandelic acid (MA) by alcohol dehydrogenase, and both are considered the major urinary metabolites of styrene and ethylbenzene exposure. ^55^ Styrene is mainly used in the manufacturing of plastics, synthetic rubbers, and resins, and is the top released carcinogen in the United States, ^56^ while ethylbenzene is found naturally in coal tar and petroleum, and used in the manufacturing of styrene. ^55^ The primary route of exposure to styrene and ethylbenzene is through inhalation, contributing up to 99% of total exposure, with 1-2% from food consumption. ^57^ We found that the ethylbenzene/styrene metabolite - PGA, but not MA was negatively associated with CACs. This could be due to the chemical structure of PGA, which is a ketone, and is therefore, more reactive than the acid metabolite MA. This could also be representative of higher ethylbenzene/styrene exposure, which would affect the ratio of PGA to MA through an increase in alcohol dehydrogenase activity. The results of our multi-VOC analysis indicate that CACs may be particularly sensitive to ethylbenzene/styrene exposures.

We found that the negative association between the N,N-DMF metabolite (AMCC) and CACs was significantly stronger in normotensive participants than those with hypertension. Previous work has shown that hypertension is associated with lower CAC levels and that hypertension is a major independent predictor of impaired endothelial progenitor cell migration. ^58^ It has also been demonstrated that hypertension accelerates EPC senescence in both clinical and experimental settings. ^59^ These findings suggest that because of the inherent susceptibility of CACs to hypertension, CACs in hypertensive participants may be more vulnerable to exposure to VOCs such as N,N-DMF. N,N-DMF is a commonly used solvent with wide industrial applications, and occupational exposure to high levels of N,N-DMF is associated with liver injury; ^60^ however, our observation that even ambient exposure to N,N-DMF is associated with vascular injury, suggests that there may be a high risk of hypertension and cardiovascular disease even with low-level exposures.

The observed association between VOC metabolites and diminished circulating angiogenic cells suggest that individuals exposed to VOCs may have a lower capacity to repair the endothelium. These effects of VOC exposure appear to be similar to those of other environmental pollutants, which are also associated with suppression of CAC levels. Previous work has shown that exposure to PM_2.5_ over a period of 24h to several weeks suppresses EPC levels, ^41, 61^ and exposure to pollutants such as particulate nickel, ^62^ acrolein ^17^ and benzene ^18^ have also been shown to reduce CAC levels. These observations suggest that pollutant exposure either depletes CAC levels in peripheral blood or interferes with the release from the bone marrow and the spleen. However, in contrast to such suppression, CAC levels are slightly elevated in individuals breathing coarse PM ^63^ or those living close to a major roadway. ^64^ This biphasic response is consistent with the dose-response relationships observed in the current study, which are indicative of increased CAC mobilization (to promote repair) at low levels of exposure, and CAC suppression at higher exposures.

Recently, we showed that living in areas of high vegetation is associated with increased levels of CD34^+^/CD45^+^/AC133^+^ CACs, but decreased CD31^+^/CD34^+^/CD45^dim^/AC133^+^ CACs, ^65^ suggesting greenness may be protective against air pollution related health effects. Thus, it seems that low levels of toxicity could lead to increased recruitment of CAC into the peripheral blood, whereas high levels of exposure could suppress CAC recruitment and/or depleting CAC by increasing utilization. The directionality of the effects of exposure on CAC levels may also depend upon locus of injury, some exposures may prevent recruitment, while others may induce injury that stimulates recruitment. Regardless, our results with VOCs suggest that exposure to these chemicals suppresses CAC levels in peripheral blood, but further studies are required to determine whether this is due to decreased recruitment or increased utilization.

We found that several VOC metabolite levels were negatively associated with angiogenic cells (CD45^dim^/CD146^+^/CD34^+^), but positively associated with hematopoietic endothelial cells (CD45^+^/CD146^+^), as evident from our environmental risk score results (Schematic 2).

**Scheme 2.**
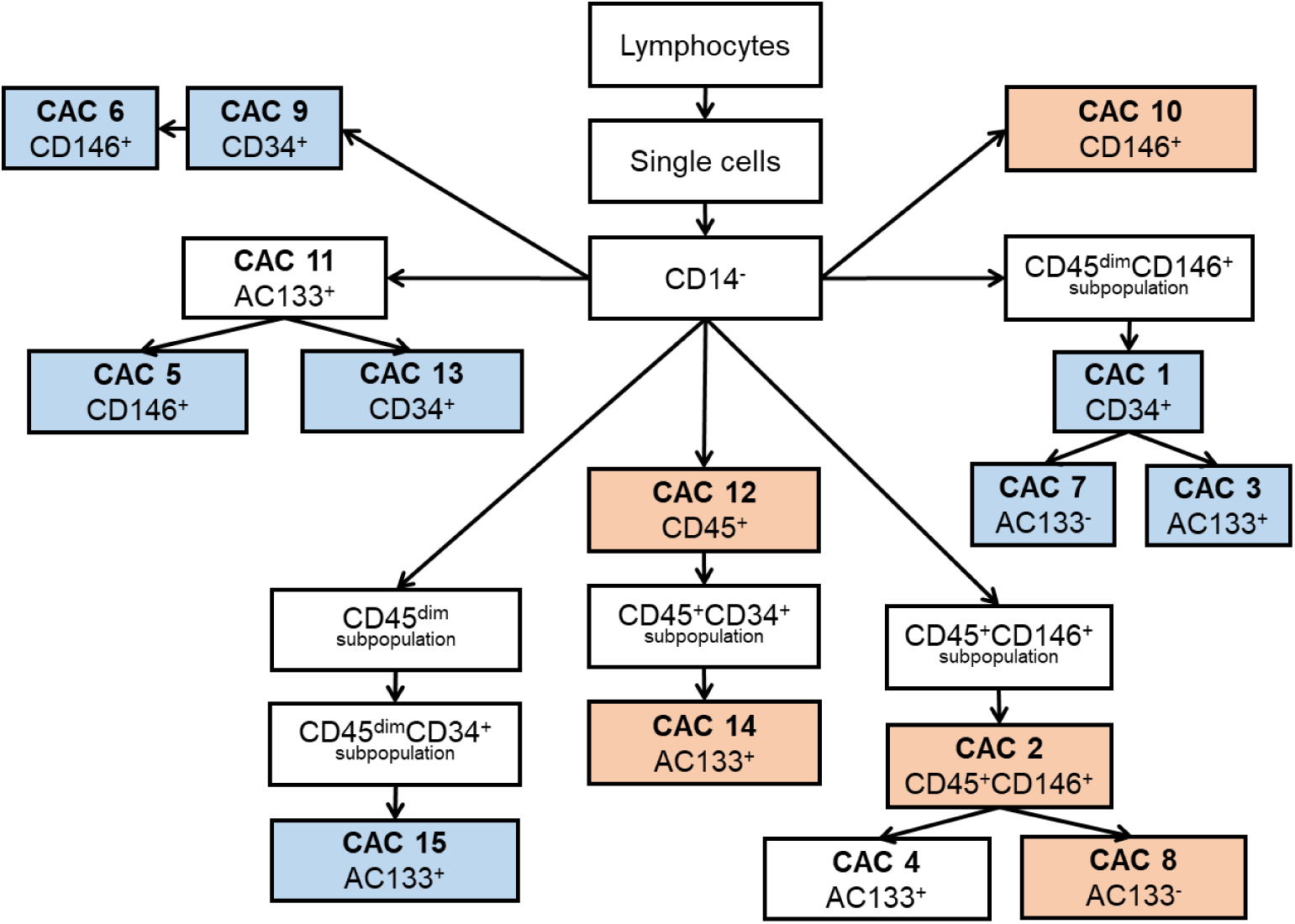
Gating Strategy for the Human Circulating Angiogenic Cells. Blue indicates negative environmental risk score; orange indicates positive environmental risk score.

In comparison with CD45^dim^ cells, CD45^+^ CACs are more primitive, with the potential to self-renew and differentiate into specialized cells that can control biological activities such as homeostasis and response to inflammation. ^66^ Additionally, the angiogenic cells are rare, and can be easily depleted when used in the repair of endothelial injury. Our exposure response curves are consistent with this notion that at low levels of VOC exposure, CD45^+^ cells have the ability to regenerate to repair endothelial injury, however at high VOC exposure, the injury surpasses the cells capacity to regenerate or the mobilization of these cells from depots such as the spleen or bone marrow is prevented. Consistent with this notion, results from our Bayesian kernel machine regression also found a nonlinear relationship with the mixture of VOCs and CAC-4 (CD45^+^/CD146^+^/AC133^+^). Further, we found that the mixture of VOC metabolites was positively associated with CD146^+^ cells. These cells are thought to be mature cells which detach from the intimal monolayer after endothelial injury. Studies have shown an increased number of CD146^+^ cells in CVD, such as unstable angina, acute myocardial infarction, stroke, and diabetes. ^67^ Positive associations could also be due to the specific VOC exposure. We found primarily positive relationships between the toluene metabolite (BMA) and CACs. This could be reflective of the low level of toxicity of toluene in comparison to other VOCs.

The major strength of this study is the use of individual level data to assess exposure. Although, measurements of the urinary metabolites do not permit source assignment, they do provide an integrated measure to total exposure of an individual whether via inhalation, ingestion, or endogenous production. Hence, in linking exposure to outcome, such measurements provide a unique advantage as exposure-response could be concurrently evaluated in the same individual. Using this approach, we were able to assess association between exposure and a sensitive and robust measure of preclinical cardiovascular injury (CACs) which is predictive of CVD events and mortality. In our analysis, we also adjusted for exposure to PM_2.5_ and were able to apply a composite VOC risk score that could be used in future studies.

Our study also has some limitations. Because of the cross-sectional nature of the study, the associations found in this study cannot address causality. Longitudinal studies are needed to determine whether VOC exposures are associated with adverse clinical outcomes, and whether these outcomes are related to changes in CAC levels. Our study also has the potential for type I errors due to testing of multiple CACs and VOC metabolites. However, due to the correlation between VOC metabolites, and limited prior research on VOC exposure and CVD, we did not adjust for multiple comparisons, to preserve our type II error, and identify potential VOC candidates for future studies. Moreover, we used spot urine collection from each individual to estimate VOC exposure. Although, it has been reported that the inter-day reproducibility for most urinary VOC metabolites is good to excellent.^68, 69^ Following exposure, many VOCs rapidly undergo biotransformation to generate metabolites that can conjugate with glutathione. Other enzymatic reactions remove glutamic acid and glycine to produce a cysteine conjugate, which is N-acetylated and excreted in the urine as mercapturic acid. ^13^ It has also been shown that smoking one cigarette, which generates high levels of VOCs, is sufficient to immediately affect the levels of CACs. ^70^ Therefore, because VOC metabolites and CAC levels were concurrently measured, it is likely that the observed association represent consequences of recent exposure. Additionally, our urinary VOC metabolites were normalized to creatinine to account for dilution differences, which could introduce error that relates to the dependence of creatinine excretions due to muscle mass, physical activity, and kidney disease. Another limitation of our study is that most participants were, by study design, of older age and at increased risk of cardiovascular disease. Thus, our sample could represent a group of susceptible individuals, which may limit generalizability of our findings.

In conclusion, we showed that metabolites of several VOCs were negatively associated with circulating levels of CD45^dim^/CD146^+^/CD34^+^ cells and the mixture of VOC metabolites was positively associated with CD146^+^ cells in the peripheral blood. These findings suggest that exposure to low ambient levels of VOCs could damage the endothelium, and deplete rare CACs or inhibit their recruitment, and thereby compromise endothelial repair. Given the global burden of CVD, and the ubiquitous distribution of VOCs, it is imperative to further assess their contribution to CVD risk. Because several commonly used products, such as paints, self-care products, and glue, are important source of VOCs, and that there are several sources of indoor emissions (compressed wood, fire-retardants), this study raises the possibility that environmental exposure to VOCs may be an important determinant of CVD risk. Our work also underscores the need for greater awareness of the potential dangers of VOC exposure and heightened vigilance against such exposures. Future longitudinal and mechanistic studies are needed to confirm current findings. If substantiated, an association between VOC exposure and CVD risk could be included in estimates of CVD risk and appropriate limits and recommendations could be developed to reduce both ambient and indoor VOC exposures.

## Data Availability

N/A

## Acknowledgements

The authors would like to thank all the partners, members, and volunteers involved in the HEAL study.

*The authors declare they have no actual or potential competing financial interests.*

## Supplemental Results

**Supplemental Table 1.**
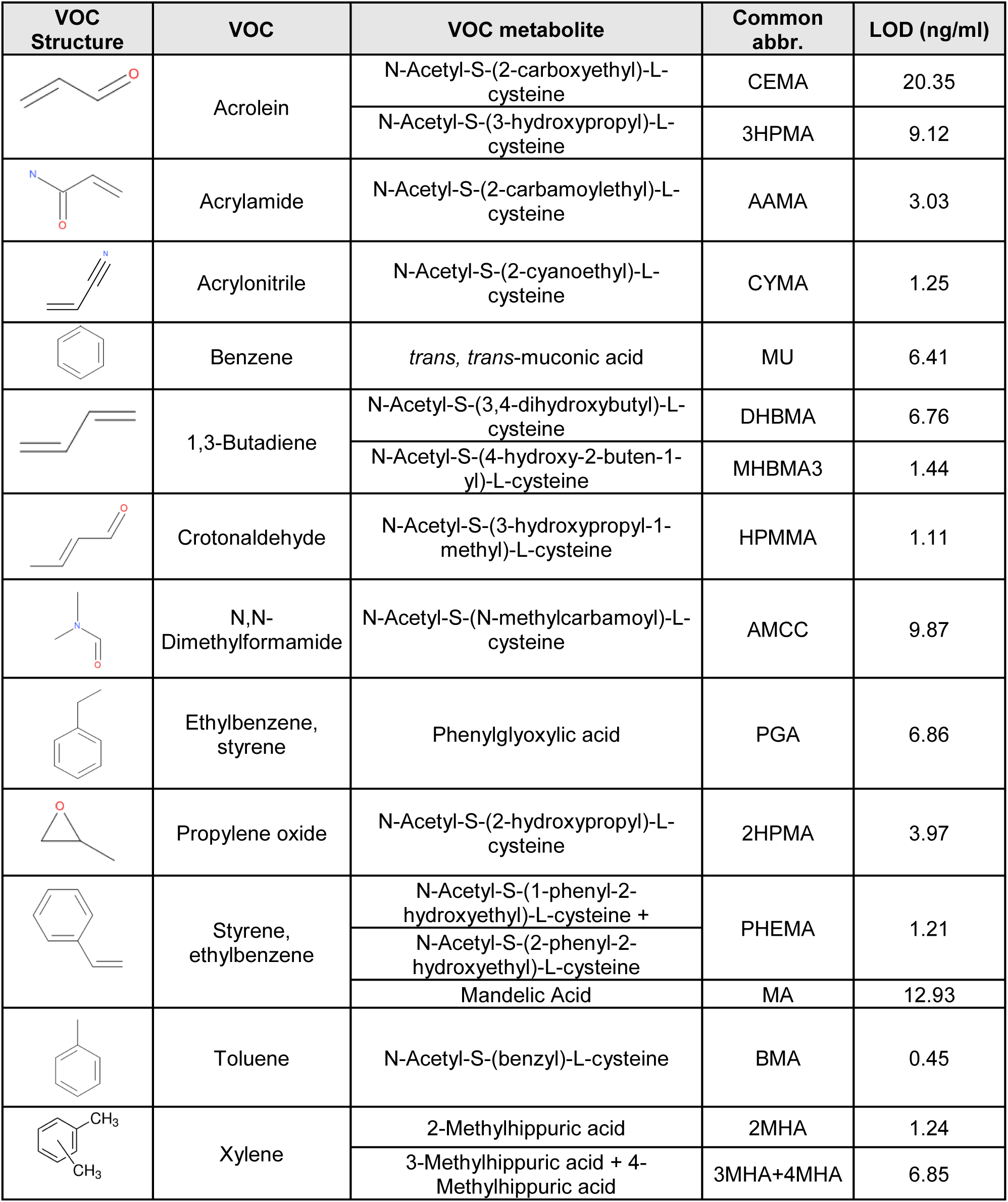
Urinary VOC metabolites and Parent Compound

**Supplemental Table 2.** Summary Statistics of urinary VOC metabolites in non-smokers for specific subgroups.

| VOC metabolite | Male | Female | White | Black | Age <55 | Age ≥55 | BMI <30 | BMI ≥30 |
| --- | --- | --- | --- | --- | --- | --- | --- | --- |
| n | 135 | 240 | 311 | 43 | 202 | 173 | 199 | 176 |
| CEMA | 104.1 ± 87.6 | 122.4 ± 130.4 | 111.2 ± 102.5 | 136.1 ± 110.7 | 102.1 ± 78.4 | 131.8 ± 148.7 | 113.1 ± 132.6 | 118.9 ± 96.8 |
| 3HPMA | 326.2 ± 269.2 | 350.2 ± 380.8 | 333.6 ± 313.3 | 330.3 ± 395.3 | 337.3 ± 325.8 | 346.5 ± 366.3 | 346.3 ± 310.9 | 336.1 ± 380.0 |
| AAMA | 73.4 ± 56.9 | 71.8 ± 64.2 | 71.2 ± 63.5 | 83.2 ± 53.5 | 72.2 ± 56.8 | 72.7 ± 66.9 | 77.9 ± 72.1 | 66.2 ± 46.3 |
| CYMA | 4.8 ± 12.3 | 6.1 ± 19.4 | **4.7 ± 15.5** | **12 ± 23.3** | 6.4 ± 17.0 | 4.8 ± 17.4 | 6.8 ± 20.6 | 4.4 ± 12.2 |
| MU | 125.8 ± 222.5 | 137.7 ± 209.4 | **146 ± 228.7** | **62.2 ± 52.1** | 126.0 ± 208.3 | 142.0 ± 220.8 | 134.8 ± 229.1 | 131.7 ± 196.2 |
| DHBMA | 348.7 ± 182.2 | 356.4 ± 172.6 | 363.6 ± 181.3 | 311.8 ± 148.3 | **328.0 ± 157.2** | **383.5 ± 191.6** | 355.0 ± 166.6 | 352.1 ± 186.3 |
| MHBMA3 | 8.4 ± 8.6 | 9.5 ± 8 | 9.3 ± 8.6 | 8.2 ± 6.1 | 9.2 ± 7.9 | 9.0 ± 8.6 | **10.2 ± 9.6** | **7.9 ± 6.1** |
| HPMMA | 286.1 ± 271.3 | 321.5 ± 324.1 | 305.1 ± 265.4 | 347.3 ± 529.9 | **275.4 ± 241.8** | **328.6 ± 267.7** | 315.1 ± 292.7 | 282.8 ± 203.8 |
| AMCC | **140 ± 99.4** | **171.9 ± 125.1** | **169.3 ± 121.1** | **118.5 ± 74.2** | **143.2 ± 104.3** | **180.5 ± 128.3** | 165.7 ± 119.9 | 154.5 ± 114.4 |
| PGA | **202.2 ± 117.2** | **239.1 ± 157.9** | 232.2 ± 151.8 | 196.3 ± 105 | 215.5 ± 149.0 | 238.0 ± 140.9 | 226.8 ± 154.1 | 224.9 ± 135.7 |
| 2HPMA | 67.7 ± 160.1 | 53.1 ± 95.8 | 60.1 ± 131.4 | 40.8 ± 31.8 | 58.1 ± 116.8 | 58.6 ± 130.0 | 67.1 ± 147.8 | 48.4 ± 86.0 |
| PHEMA | **2.3 ± 5.4** | **2.2 ± 3** | 2.3 ± 4.4 | 1.7 ± 1.2 | 2.1 ± 3.9 | 2.3 ± 4.2 | **2.2 ± 3.9** | **2.2 ± 4.2** |
| MA | **206.3 ± 124.1** | **247.7 ± 183.3** | **243.7 ± 175.6** | **177.8 ± 91.8** | **210.7 ± 139.5** | **258.7 ± 188.7** | 231.4 ± 156.0 | 234.5 ± 176.0 |
| BMA | **10.7 ± 12.4** | **12 ± 11.9** | 11.4 ± 12.1 | 11.6 ± 11.8 | **10.5 ± 11.9** | **12.7 ± 12.2** | **12.2 ± 12.8** | **10.7 ± 11.1** |
| 2MHA | 26.3 ± 37.1 | 36.5 ± 89.8 | 33.8 ± 78.7 | 25.5 ± 32.6 | 27.6 ± 50.4 | 32.9 ± 56.7 | **37.9 ± 63.7** | **21.2 ± 36.8** |
| 3MHA+4MHA | 189 ± 313.1 | 220.5 ± 454.3 | 213 ± 436 | 193.8 ± 205.9 | **179.8 ± 310.6** | **212.5 ± 293.9** | 227.4 ± 367.2 | 158.4 ± 204.2 |

Supplemental Table 2 continued. Summary Statistics of urinary VOC metabolites in non-smokers for specific subgroups.
| VOC metabolite | Non-Diabetic | Diabetic | Normotensive | Hypertensive | Low PM<sub>2.5</sub> | High PM<sub>2.5</sub> |
| --- | --- | --- | --- | --- | --- | --- |
| n | 310 | 65 | 219 | 156 | 189 | 186 |
| CEMA | 116.8 ± 123.3 | 111.2 ± 81 | 104.7 ± 85.1 | 131.4 ± 149.8 | 117.0 ± 92.4 | 114.6 ± 137.9 |
| 3HPMA | 338.7 ± 341.3 | 355 ± 362.4 | 327.9 ± 315.8 | 360.6 ± 381.7 | 371.4 ± 411.7 | 311.2 ± 257.0 |
| AAMA | 74.6 ± 64.3 | 62.2 ± 45.5 | 70.7 ± 55.6 | 74.9 ± 69.3 | 65.0 ± 41.6 | 80.0 ± 76.1 |
| CYMA | 6.1 ± 18.5 | 3.6 ± 8.7 | 5.7 ± 15.9 | 5.6 ± 18.9 | 4.4 ± 12.3 | 7.0 ± 21.0 |
| MU | **128.1 ± 213.7** | **158.4 ± 215.3** | **127.5 ± 227.6** | **141.6 ± 193.7** | 142.7 ± 257.3 | 123.9 ± 158.5 |
| DHBMA | 354.3 ± 172.8 | 350.4 ± 191.2 | 347.4 ± 164.4 | 362.4 ± 191.2 | **368.6 ± 182.0** | **338.5 ± 168.6** |
| MHBMA3 | 9.3 ± 8.6 | 8.3 ± 6.2 | 8.9 ± 7.7 | 9.5 ± 8.9 | 9.1 ± 8.4 | 9.2 ± 8.1 |
| HPMMA | 300.9 ± 260.8 | 346 ± 466.9 | 277.8 ± 228 | 352.1 ± 387.2 | 308.9 ± 278.3 | 290.9 ± 229.5 |
| AMCC | 157.3 ± 108.6 | 175.1 ± 152.4 | 154.8 ± 109.2 | 168.3 ± 127.8 | 153.5 ± 102.8 | 167.4 ± 130.4 |
| PGA | **219.3 ± 143.1** | **257.2 ± 154** | 222.7 ± 156.8 | 230.3 ± 128.6 | **208.3 ± 133.0** | **243.7 ± 155.6** |
| 2HPMA | 62 ± 132.5 | 40.9 ± 55.8 | **66.1 ± 137** | **47.4 ± 99.3** | 64.8 ± 144.2 | 51.8 ± 96.5 |
| PHEMA | 2.3 ± 4.4 | 1.8 ± 1.9 | 2.3 ± 4.6 | 2.1 ± 3.2 | 2.1 ± 3.6 | 2.3 ± 4.5 |
| MA | 228.5 ± 155.5 | 253.4 ± 206.9 | 223.9 ± 154.4 | 245.3 ± 179.6 | 217.1 ± 146.2 | 248.9 ± 182.1 |
| BMA | 12 ± 12.7 | 9.4 ± 8.1 | 11.7 ± 12.7 | 11.3 ± 11.1 | 10.8 ± 11.4 | 12.3 ± 12.7 |
| 2MHA | 35 ± 80 | 22.5 ± 46.6 | 37.4 ± 91.6 | 26.5 ± 43.2 | 32.8 ± 64.7 | 27.3 ± 38.8 |
| 3MHA+4MHA | 223.7 ± 445.8 | 140.2 ± 115.9 | 226.4 ± 480 | 185.2 ± 282.4 | 211.2 ± 368.4 | 178.4 ± 218.4 |

Values represent Mean ± SD. Units: µg/g creatinine. Bold values represent significant difference between specific subgroups, based on T-test of log-transformed VOC metabolites. Low PM<sub>2.5</sub> ranges from 1.2-7.8 µg/m<sup>3</sup>. High PM<sub>2.5</sub> ranges from 8.0-17.7 µg/m<sup>3</sup>.

**Supplemental Figure 1.**
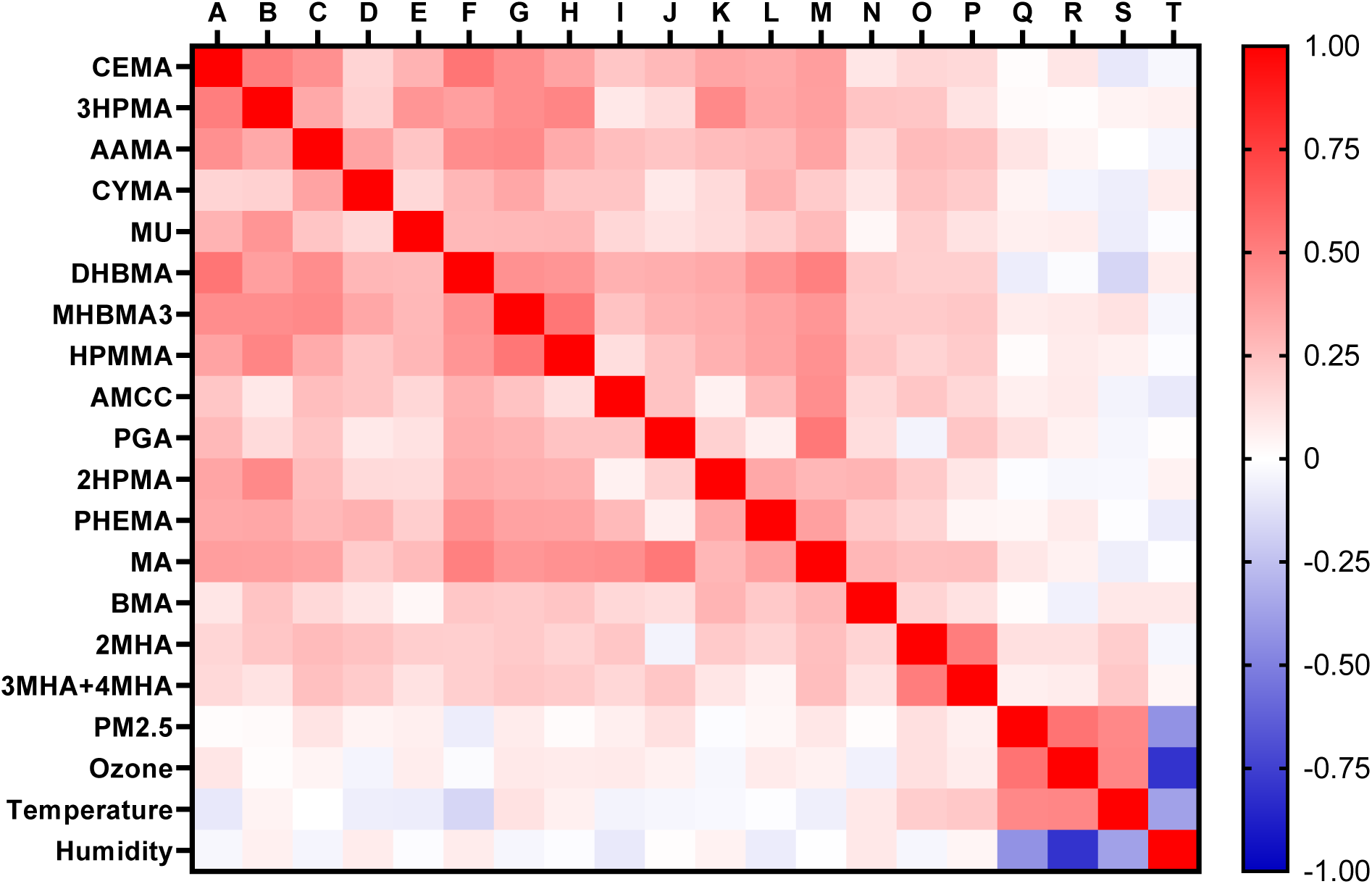
Pearson correlation coefficients between log-transformed urinary VOC metabolites and ambient pollutant and meteorological data (non-smokers; n=375).

**Supplemental Figure 2.**
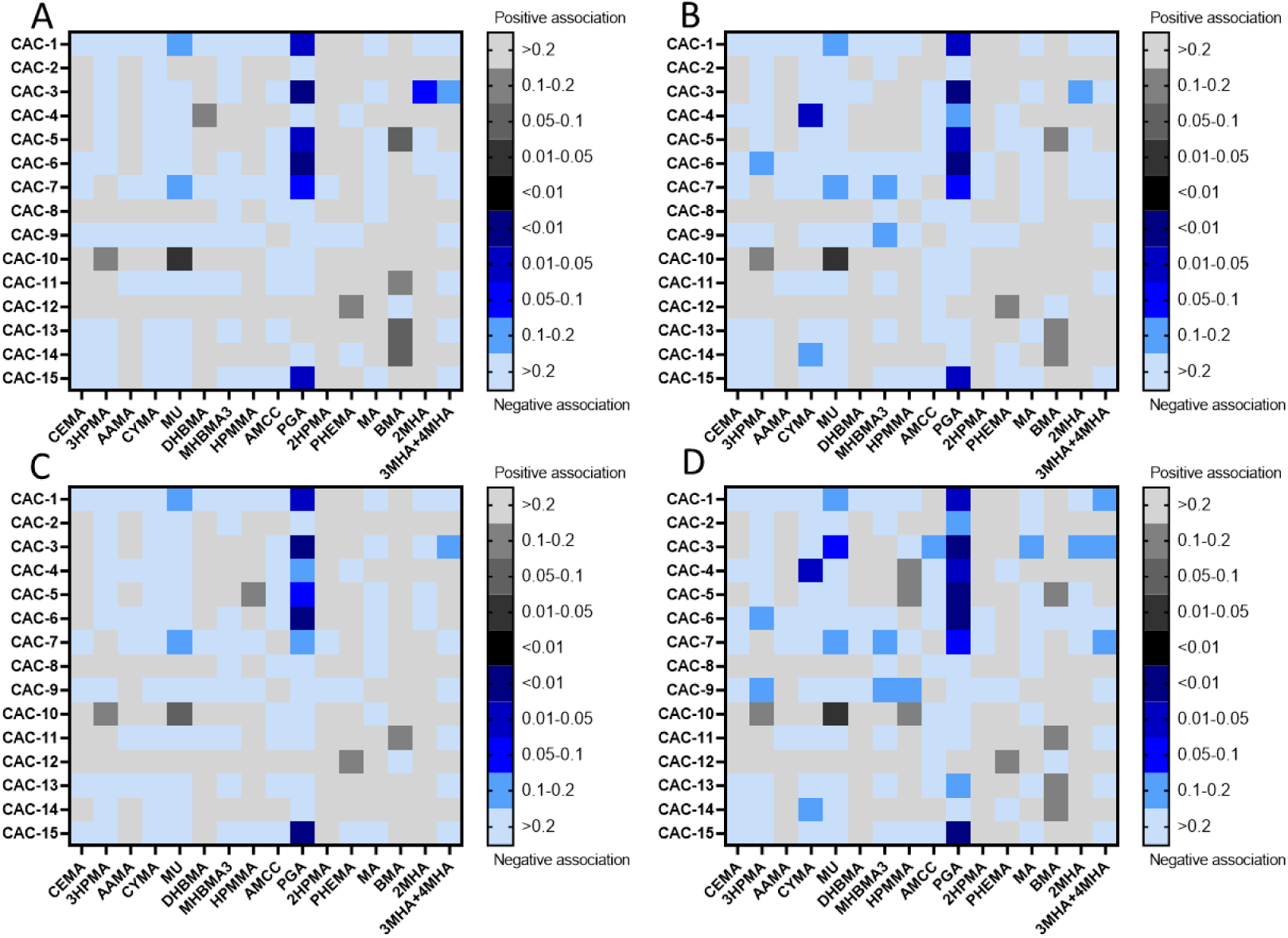
Heat map of pair-wise associations between the urinary metabolites of VOCs and circulating levels of CACs. False Discovery rate adjusted p-values. Generalized linear models (non-smokers; n=375) were adjusted for age, sex, race, BMI, diabetes, hypertension and: daily PM_2.5_ + Temperature (A); daily PM_2.5_ + Humidity (B); Ozone + Temperature (C); Ozone + Humidity (D). Black grids represent positive associations and blue grids represent negative associations. The intensity of colors represent p-values, with darker grids indicating smaller p-values.

